# Annual prevalence of non-communicable diseases and identification of vulnerable populations following the Fukushima disaster and COVID-19 pandemic

**DOI:** 10.1101/2022.05.18.22275293

**Authors:** Michio Murakami, Shuhei Nomura

## Abstract

Disasters, pandemics, and their response measures can have secondary effects on the physical and psychological health of affected populations. Identifying populations vulnerable to these effects is beneficial for promoting effective health and prevention strategies. Using health insurance receipt data from 2009 to 2020, we assessed changes in prevalence of major non-communicable diseases (NCDs), including hypertension, hyperlipidemia, diabetes, and mental disorders, among affected populations before and after the Fukushima disaster and coronavirus disease (COVID-19) outbreak in Japan. Furthermore, age and sex groups with the largest increases in prevalence after these events were identified. The participants of this study were members of the Employees’ Health Insurance scheme, including employees of companies and their dependent family members. The dataset was provided by JMDC Inc. The annual age-adjusted prevalence of each disease was used to calculate the ratio of disease prevalence before and after the events. After the Fukushima disaster, hypertension, hyperlipidemia, and diabetes generally increased over a 9-year period in Fukushima Prefecture. The increase in the prevalence rate of these three NCDs and mental disorders were the highest among females aged 40–74 years compared to males and the other age groups. The prevalence of all four diseases increased after the COVID-19 outbreak in Japan, with marked increase in males aged 0–39 years. Populations that have experienced secondary health effects such as NCDs are unique to each disaster or pandemic, and it is important to provide tailor-made public health support among populations in accordance to the type of disasters and pandemic.

**Highlights:**

- We assessed secondary health effects of Fukushima disaster and COVID-19 pandemic
- Non-communicable diseases increased after the disaster and COVID-19 pandemic
- The increase rates were higher among females aged 40–74 years after the disaster
- The increase rates were higher among males aged 0–39 years after COVID-19 pandemic
- It is important to provide tailor-made public health support among populations

## 1. Introduction

Disasters, pandemics, and their countermeasures impact the lives of affected people with secondary effects on their physical and psychological health. Major non-communicable diseases (NCDs), such as hypertension, hyperlipidemia, diabetes, and mental disorders, have been reported to increase among people who have experienced natural or man-made disasters, such as the Chornobyl nuclear power plant accident in 1986 and Hurricane Katrina in 2005 [1, 2]. Several studies have also revealed that the Great East Japan Earthquake and the Fukushima Daiichi Nuclear Power Station accident in March 2011 (hereafter referred to as the “Fukushima disaster”) also caused an increase in NCDs among residents who evacuated for short to long periods [3–5]. In addition, long-term deterioration of mental health, including increases in psychological distress and suicide rates, has been observed, especially among residents affected by the Fukushima disaster, although there is a trend toward improvement over time [6, 7]. Furthermore, in Minamisoma City, located within 10–40 km of the nuclear power station, a significant increase in the age-adjusted prevalence of diabetes was observed among both those who did not evacuate and those who did, during a 7-year period following the disaster [8]. High psychological distress has also been reported among non-evacuees in host communities where evacuees were accommodated [9]. Changes in living conditions due to evacuation after a disaster as well as various socioeconomic changes in the surrounding communities may cause secondary physical and psychological health effects to evacuees and non-evacuees. However, the spatial and temporal extent of post-disaster secondary health effects have not been established, as there are only a limited number of long-term studies on both evacuees and non-evacuees over a wide area.

Following the emergence and worldwide spread of coronavirus disease (COVID-19), behavioral restrictions such as lockdowns or stay-at-home measures, have been implemented in various countries, including Japan, to curb infection spread. During the pandemic, some studies have shown improvement in lifestyle, dietary, and drinking behaviors [10–12]; however, increases in body weight and worsening of metabolic biomarkers, such as glycemic values, among specific populations have also been reported [13, 14]. Furthermore, there has been a significant increase in mental disorders among the global population, especially in females and younger age groups [15]. Similarities and differences in secondary health effects among multiple disasters and pandemics remain unclear due to limited analyses of data obtained before and after such events. Furthermore, identifying the age and sex groups in which NCDs occurred can provide beneficial information to promote effective support for vulnerable populations regarding the secondary health effects following disasters and pandemics.

We used health insurance receipt data to assess changes in the prevalence of hypertension, hyperlipidemia, diabetes, and mental disorders in the population affected by the Fukushima disaster and COVID-19 in Japan. First, we investigated how the prevalence of these diseases changed over 9 years from January 2009 to December 2020 following the Fukushima disaster. The Fukushima, neighboring prefectures of Iwate, and Miyagi prefectures, as well as the whole of Japan, and three major regions within Fukushima Prefecture were considered to assess the wide-area secondary health effects caused by the disaster. We then analyzed the prevalence of these diseases in the Japan and Fukushima Prefecture by age and sex to identify the populations that were the most significantly affected. Next, we examined the annual changes in the prevalence of these diseases across Japan before and after the COVID-19 outbreak and identified the most significantly affected age and sex groups. We then analyzed the associations between population density or COVID-19 infection rates and changes in the prevalence of these diseases by prefecture to determine prefectural characteristics that are susceptible to these secondary health effects.

## 2. Methods

### 2.1. Ethics

This study was approved by the Osaka University Research Ethics Committee (approval number Ci21002).

### 2.2. Fukushima disaster

The reported number of people directly killed or missing due to the Great East Japan Earthquake and Tsunami (GJET) on March 11, 2011 was 18,423 (at the end of February 2022 [16]). The reported number of disaster-related deaths caused indirectly by the GJET (i.e., secondary victims) was 3,784 (at the end of September 2021) [17]. Of the 18,423 people, 5,785 people in Iwate Prefecture, 10,757 in Miyagi Prefecture, and 1,810 in Fukushima Prefecture were killed or missing. There were 470 disaster-related deaths in Iwate Prefecture, 929 in Miyagi Prefecture, and 2,329 in Fukushima Prefecture. The number of people living in evacuation shelters immediately after the GJET was estimated to be more than 450,000 nationwide, including more than 400,000 in the Iwate, Miyagi, and Fukushima prefectures [18].

Following the Fukushima disaster, mandatory evacuation orders were issued to residents within a 20- km radius around the Fukushima Daiichi Nuclear Power Station and a 10-km radius around the Daini Nuclear Power Station on March 12, 2011, followed by evacuation orders on April 22, 2011 for areas where an additional exposure level would reach 20 mSv/y. Evacuation orders were issued for Hirono Town, Naraha Town, Tomioka Town, Kawauchi Village, Okuma Town, Futaba Town, Namie Town, Katsurao Village, Iitate Village, Minamisoma City, Tamura City, Kawamata Town, and the hotspot areas in Date City. Of these, Tamura City, Kawamata City, and Date City are in Nakadori (central area), whereas the other municipalities are located in Hamadori (coastal area). It was estimated that more than 140,000 people were evacuated from these radius [19]. The affected people were evacuated to municipalities in Fukushima Prefecture outside the evacuation areas, such as Iwaki City (Hamadori), as well as to areas outside Fukushima Prefecture.

In 2012, the evacuation order was lifted in Kawauchi Village and Hirono Town, where the evacuation was decided by each municipality soon after the disaster. Furthermore, since 2014, the government has gradually lifted the evacuation order, and by the end of 2019, the evacuation order was lifted for the following municipalities: Tamura City, Kawauchi Village, Naraha Town, Katsurao Village, Minamisoma City, Iitate Village, Kawamata Town, Namie Town, Tomioka Town, and Okuma Town, excluding the difficult-to-return areas where radiation levels remain high located around the Fukushima Daiichi Nuclear Power Station.

### 2.3 COVID-19

The World Health Organization (WHO) declared COVID-19 a pandemic on March 11, 2020 [20]. In Japan, a declaration of the state of emergency was issued in seven prefectures on April 7, 2020 and extended to all prefectures on April 16, 2020. The state of emergency included requests to stay at home, instruction for schools to close temporarily, closure of businesses and stores that are not essential to daily life, and cancellation or postponement of events and exhibitions, but they were not mandatory. The state of emergency ended in 39 prefectures on May 14, 2020, in three more prefectures on May 21, 2020, and in all prefectures on May 25, 2020. This was the only emergency declaration for 2020, and the reported number of COVID-19 positive cases in Japan in 2020 was 234,109 [21].

### 2.4. Datasets and target population

In this study, we used health insurance receipt data provided by JMDC Inc. The target population covered in the JMDC data were employees of companies and their dependent family members who belonged to Employees’ Health Insurance (EHI) scheme (29 million people nationwide as of 2016), of which the JMDC covered 9 million of the population (as of 2022). JMDC Inc. contracts various health insurers under the EHI scheme across Japan to collect insurance claims, containing completely anonymized receipt information. Japan’s insurance system is divided into three main schemes, and all persons, regardless of citizenship, who have been recognized as having resided in Japan for more than three months are obliged to join one of them. In addition to the EHI scheme, which is the focus of this study, the insurance system includes National Health Insurance (NHI), which targets individuals who do not belong to the EHI scheme, including farmers, freelancers, non-regular employees, retired company employees, etc., and long-term care insurance (LTCI), which includes individuals aged 75 and above.

We used data on the annual number of patients for each targeted disease and other diseases (i.e., all patients visiting the medical institution regardless of their diagnosis) and the estimated prevalence by age and sex groups in each prefecture or municipality from January 2009 to December 2020. The prefectures and municipalities were based on the location of the medical institution. The target diseases included hypertension, hyperlipidemia, diabetes, and mental disorders, based on the ICD 10 classification: I10-I15, E78, E10-E14, and F00-F99, respectively. The diseases were diagnosed by a physician, with no distinction made between primary and secondary diseases. The total number of patients in the JMDC database for hypertension, hyperlipidemia, diabetes, mental disorders, and other disease in Japan from 2009 to 2020 is provided in Table S1.

The prevalence (*Pd,t,r,a,s*) of the target disease (*d*) in each prefecture or municipality (*r*) by age (*a*:0– 19, 20–39, 40–64, and 65–74 years) and sex (*s*) groups for each year (*t*) were calculated according to eqs.1 and 2.

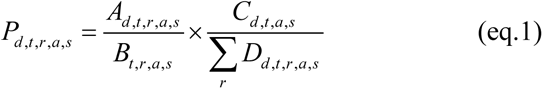

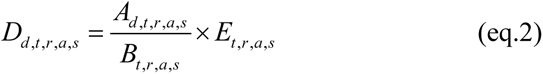

Where *Ad,t,r,a,s* is the actual number of patients with the target disease by each prefecture or municipality among the target age and sex population in the target year, *Bt,r,a,s* is the actual number of patients who visited the medical institution regardless of their diagnosis in each prefecture or municipality among the target age and sex population in the target year, *Cd,t,a,s* is the estimated number of patients nationwide among the target age and sex population in the target year and was estimated by multiplying the prevalence of each disease (the ratio of the number of patients to the number of people in the mother population covered by JMDC Inc.) by age (1-year interval) and sex groups and by the national population, *Dd,t,r,a,s* is the estimated number of patients with the target disease in each prefecture and municipality in the target age and sex population in the target year, and *Et,r,a,s* is the number of people in each prefecture and municipality in the target age and sex population in the target year.

The Basic Resident Register data was used for to determine the number of people in the prefectures for each year [22]. For the municipal population data, the national census performed in 2010, 2015, and 2020 was used for the years 2009–2010, 2011–2015, and 2016–2020, respectively [23]. Selection bias among the target population was corrected by *Cd,t,a,s* and *Dd,t,r,a,s*, as described in eq.1.

Furthermore, using the commonly used 1985 model population as a reference population (i.e., distribution of the population by age and sex groups in 1985) [24], age-adjusted prevalence was calculated for the 0–74, 0–39, and 40–74 years of age groups for males, females, and both males and females.

Diagnostic guidelines have been revised for various diseases, including the establishment of borderline hyperLDL-cholesterolemia in 2012 and the diagnosis of diabetes using HbA1c in 2010. Although these revisions may have impacted the prevalence, the bias was reduced by standardizing the assessment of the impact before and after the Fukushima disaster by normalizing the prevalence with that for the whole of Japan (see “2.5. Analysis” for details).

### 2.5. Analysis

Two analyses were performed in this study. First, to evaluate changes in prevalence after the Fukushima disaster, we considered the whole of Japan, Iwate, Miyagi, and Fukushima prefectures, as well as three regions within Fukushima Prefecture (i.e., Hamadori, Nakadori, and Aizu (mountainous area); Table S2). These three regions, which are closer to the nuclear power station in the order Hamadori, Nakadori, and Aizu, divide Fukushima Prefecture into three parts from east to west. In other words, Hamadori was the region most affected by the disaster, and Aizu was the region least affected among regions in the prefecture. Prevalence of targeted diseases in the Hamadori, Nakadori, and Aizu regions was calculated by weighting data by municipality based on *Bt,r,a,s*. The arithmetic means and standard errors of the prevalence for each of the four time periods, such as 2009–2010, 2011–2013, 2014–2016, and 2017–2019, was calculated, and the prevalence for each time period was assumed to follow a normal distribution. To ensure that the prevalence was within the range of 0–1, two-tailed censoring was performed so that the arithmetic mean was at the center of the distribution. Monte Carlo simulations were performed with 10,000 iterations to determine the ratio or difference in prevalence (males and females, 0–74 years) between 2011–2013, 2014–2016, or 2017–2019 and 2009–2010 in the whole of Japan, Iwate, Miyagi, Fukushima prefectures, and Hamadori, Nakadori, and Aizu regions. These prevalence ratios and differences were further normalized to the values for the whole of Japan. Similarly, prevalence ratios and differences by age and sex groups were calculated for the whole of Japan and Fukushima Prefecture.

Second, to assess changes in the prevalence after the COVID-19 outbreak, data from 2015 to 2020 were used (Table S3). For simplicity, we defined 2020 data as the period after the COVID-19 outbreak. We calculated the ratios and differences between the prevalence in 2020 (the period after the outbreak) and the prevalence in 2020 estimated from the 2015–2019 dataset. A linear regression equation was modeled using the dataset as a baseline to estimate the hypothetical 2020 prevalence in the absence of COVID-19. Based on the values and their 95% confidence intervals, calculations of the ratios and differences between the measured and estimated prevalence were performed with Monte Carlo simulations in 10,000 runs, assuming that the prevalence followed a normal distribution; two-tailed censoring was performed so that the prevalence fell within the range of 0–1, and the arithmetic mean was at the center of the distribution. The prevalence ratios and differences were estimated for age and sex groups in the whole of Japan: males and females (0–74 years), males and females (0–39 years), males and females (40–74 years), males (0–74 years), males (0–39 years), males (40–74 years), males (0–19 years), males (20–39 years), males (40–64 years), males (65–74 years), females (0–74 years), females (0–39 years), females (40–74 years), females (0–19 years), females (20–39 years), females (40–64 years), and females (65–74 years). Furthermore, the prevalence ratios and differences in 47 prefectures were calculated for males (0–39 years), the vulnerable population group identified in the above analysis, to assess the correlation with population density [22, 25] or the reported number of COVID-19 infection cases per 100,000 people (COVID-19 infection rate) in 2020 [21] (Table S4).

Spearman’s rank correlation coefficient was estimated.

Oracle Crystal Ball (Version 11.1.2.4.900) and IBM SPSS Statistics (Version 28) were used for the analysis.

## 3. Results

### 3.1. Age-adjusted prevalence over time

Figure 1 shows the annual changes in age-adjusted prevalence (male and female, 0–74 years) of NCDs from 2009 to 2020 in the whole of Japan; Iwate, Miyagi, and Fukushima prefectures; and Hamadori, Nakadori, and Aizu regions. The age-adjusted prevalence of hypertension, hyperlipidemia, diabetes, and mental disorders in the whole of Japan was 0.102, 0.107, 0.061, and 0.075 in 2009 and 0.113, 0.121, 0.068, and 0.098 in 2020, respectively. The age-adjusted prevalence of hypertension, hyperlipidemia, diabetes, and mental disorders in Fukushima Prefecture was 0.106, 0.098, 0.044, and 0.072 in 2009 and 0.117, 0.119, 0.056, and 0.082 in 2020, respectively.

**Figure 1.**
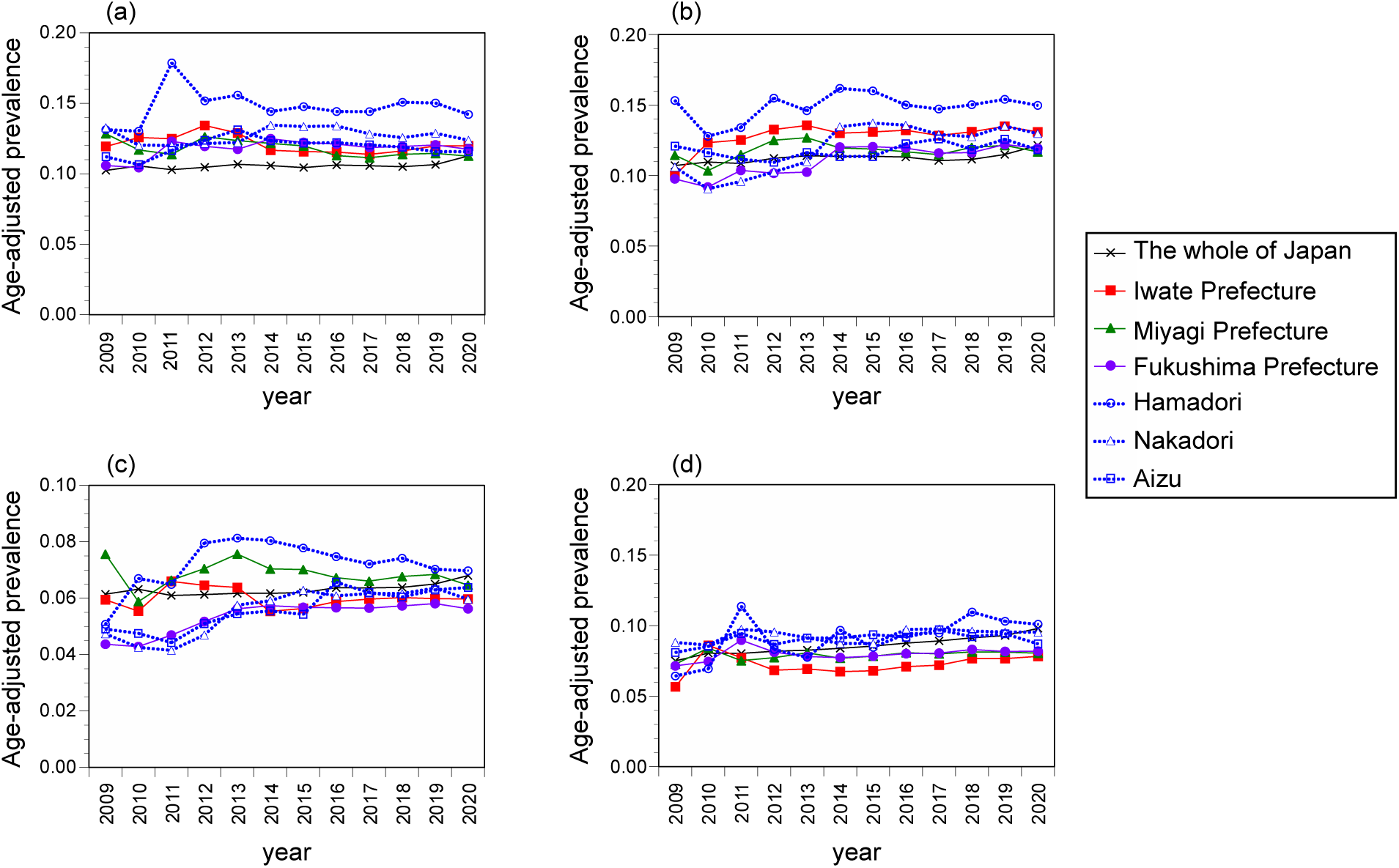
Annual age-adjusted prevalence of hypertension, hyperlipidemia, diabetes, and mental disorders by areas (males and females, 0–74 years). (a) Hypertension, (b) hyperlipidemia, (c) diabetes, (d) mental disorders.

Age-adjusted prevalence by age and sex groups for each disease in the whole of Japan and Fukushima Prefecture from 2009 to 2020 is shown in Figures 2 and 3. Age-adjusted prevalence of hypertension, hyperlipidemia, and diabetes were generally higher in males than in females in the 40–74 years age group, and the differences tended to increase over time in the whole of Japan. In contrast, mental disorders were higher in females than in males in the 40–74 years age group, and the prevalence in the 0–39 years age group has been increasing in recent years. In Fukushima Prefecture, mental disorders showed an increasing trend in 2011–2012 for both age and sex groups, followed by a decreasing trend.

**Figure 2.**
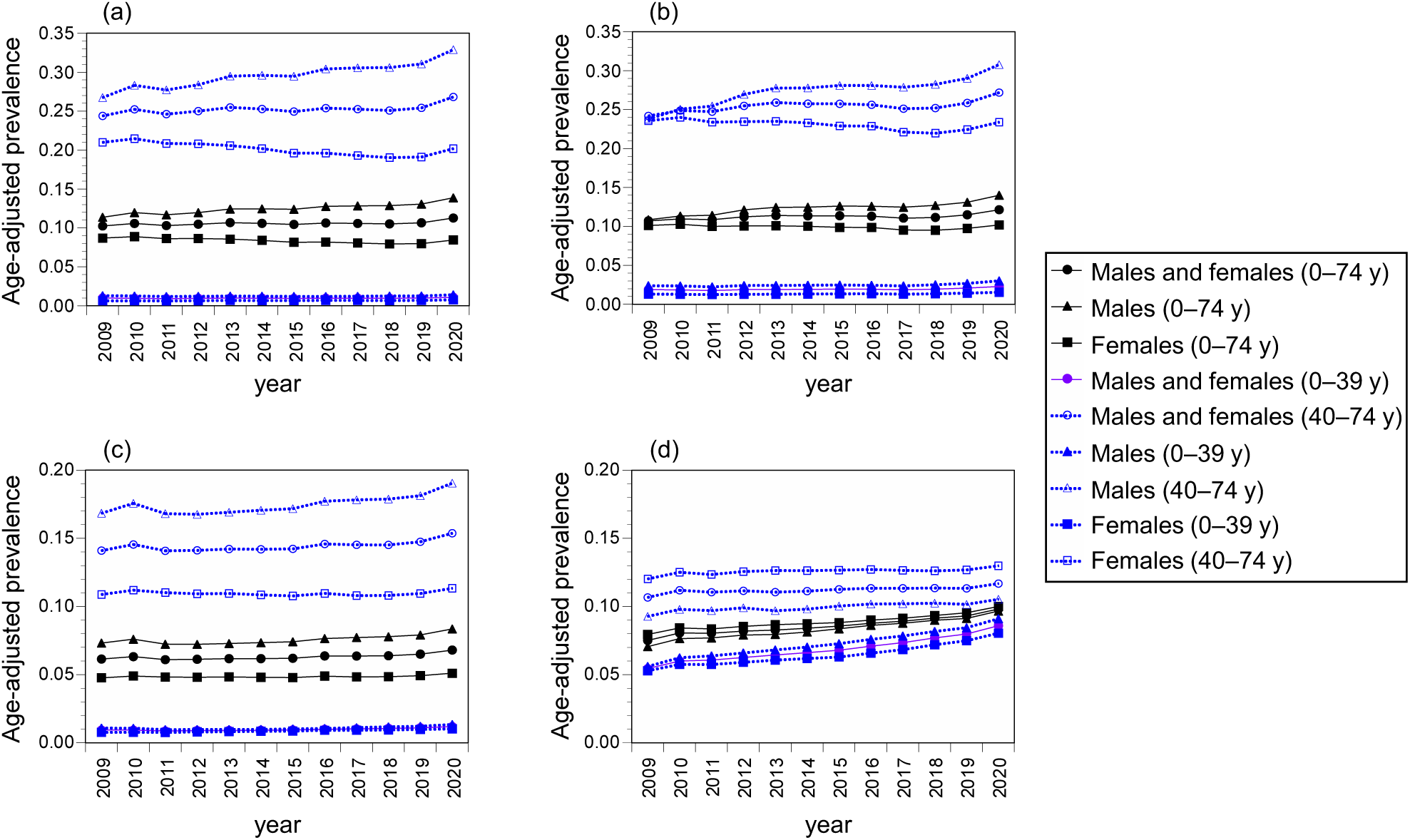
Annual age-adjusted prevalence of hypertension, hyperlipidemia, diabetes, and mental disorders by age group and sexes (the whole of Japan). (a) Hypertension, (b) hyperlipidemia, (c) diabetes, (d) mental disorders.

**Figure 3.**
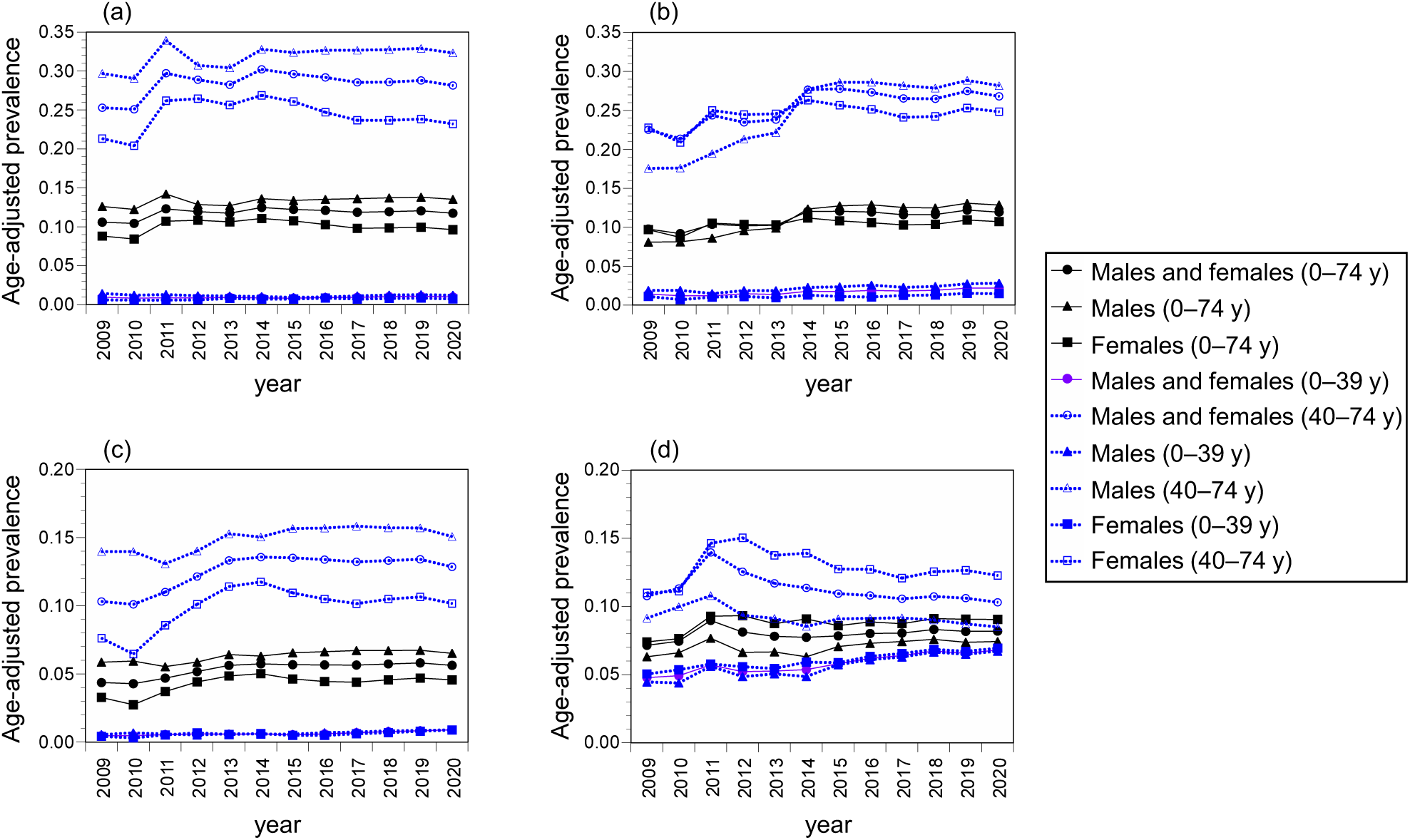
Annual age-adjusted prevalence of hypertension, hyperlipidemia, diabetes, and mental disorders by age groups and sexes (Fukushima Prefecture). (a) Hypertension, (b) hyperlipidemia, (c) diabetes, (d) mental disorders.

### 3.2. Age-adjusted prevalence before and after the Fukushima disaster

Table 1 shows the ratios of age-adjusted prevalence of NCDs in the 0–74 years group in the whole of Japan; Iwate, Miyagi, and Fukushima prefectures; and Hamadori, Nakadori, and Aizu regions before and after the Fukushima disaster. These differences are listed in Table S5. The table also shows the values normalized to those for the whole of Japan. After the Fukushima disaster, the increases in the prevalence of hypertension, hyperlipidemia, and diabetes were significantly higher among the Fukushima Prefecture than among the whole of Japan in all the three time periods including 2011– 2013, 2014–2016, and 2017–2019 (excluding hyperlipidemia in 2011–2013). The prevalence ratios in 2011–2013 were 1.133 (95% uncertainty interval (UI): 1.081–1.188) for hypertension, 1.053 (0.980–1.132) for hyperlipidemia, 1.212 (1.082–1.344) for diabetes, and 1.084 (0.961–1.212) for mental disorders. Furthermore, in Hamadori, the prevalence of hypertension, diabetes, and mental disorders generally increased over the 9-year period while in Nakadori, significant increases in the prevalence of hyperlipidemia and diabetes were observed in 2014–2016 and 2017–2019. In Aizu, there were also significant increases in the prevalence of hypertension in 2011–2013 and 2014–2016 and of diabetes in 2014–2016 and 2017–2019. In contrast, a slight downward trend was observed for hyperlipidemia in 2011–2013. No significant increases or decreases were observed for these four diseases in Iwate and Miyagi prefectures (excluding hypertension in 2014–2016 and diabetes in 2011–2013 in Iwate Prefecture).

**Table 1.**
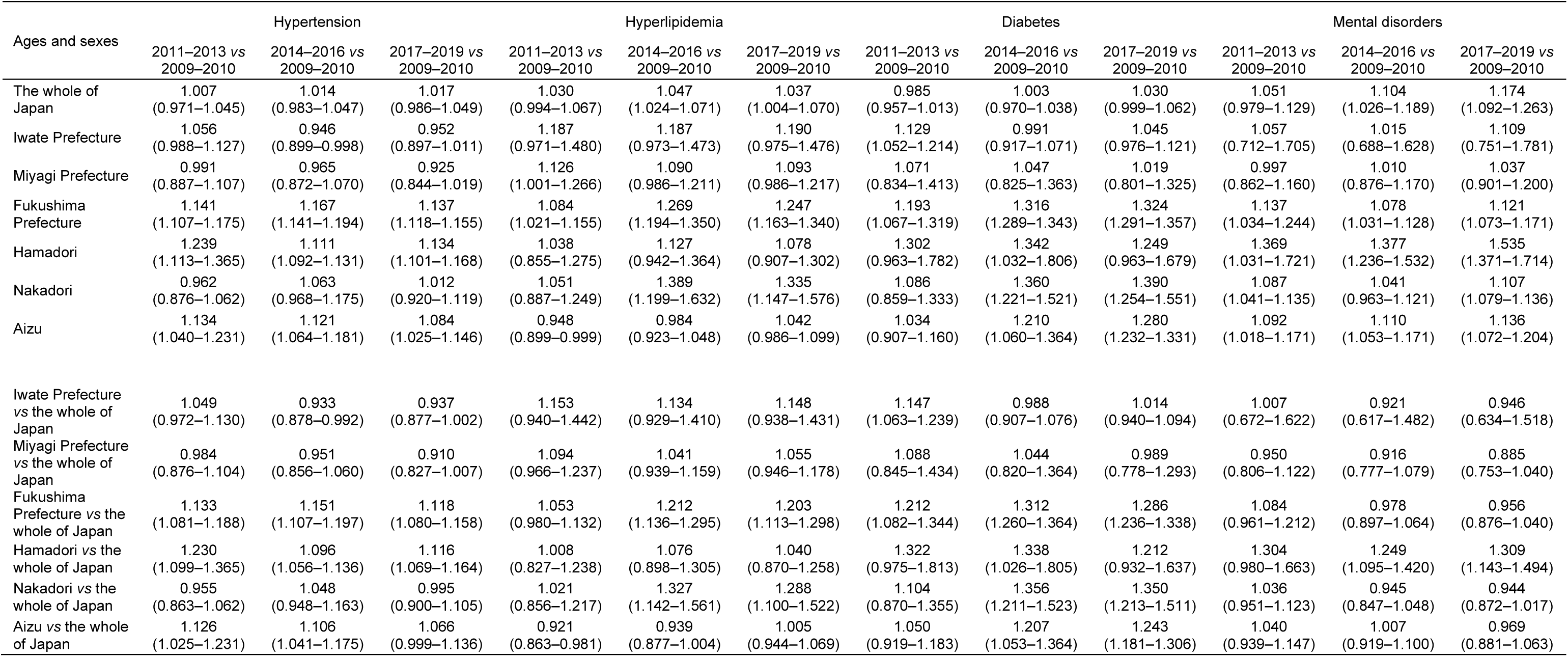
Ratio of age-adjusted prevalence in three time periods to that in 2009–2010 (males and females, 0–74 years). Arithmetic mean (95% uncertainty interval).

Regarding age and sex groups, there was a significant increase in the prevalence of these four diseases in Fukushima Prefecture during all the three time periods in females aged 40–74 years: ratios in 2011– 2013 were 1.280 (95%UI:1.214–1.350) for hypertension, 1.149 (1.055–1.257) for hyperlipidemia, 1.441 (1.139–1.806) for diabetes, and 1.283 (1.199–1.371) for mental disorders (Table 2, ratios and Table S6, difference). An increase in the prevalence of hypertension and diabetes was observed in females aged 0–39 years. In contrast, among males aged 40–74 years, the prevalence of hyperlipidemia and diabetes increased significantly in 2014–2016 and 2017–2019, whereas the prevalence of mental disorders decreased in 2014–2016 and 2017–2019. Among males aged 0–39 years, there was an increase in the prevalence of hyperlipidemia in 2014–2016 and 2017–2019, whereas there was no significant change in the prevalence of other diseases (except hypertension in 2014–2016).

**Table 2.** Ratio of age-adjusted prevalence in three time periods to that in 2009–2010 by age groups and sex (Fukushima Prefecture *vs* the whole of Japan). Arithmetic mean (95% uncertainty interval).

| Ages and sexes | Hypertension |  |  | Hyperlipidemia |  |  | Diabetes |  |  | Mental disorders |  |  |
| --- | --- | --- | --- | --- | --- | --- | --- | --- | --- | --- | --- | --- |
|  | 2011–2013 vs<br>2009–2010 | 2014–2016 vs<br>2009–2010 | 2017–2019 vs<br>2009–2010 | 2011–2013 vs<br>2009–2010 | 2014–2016 vs<br>2009–2010 | 2017–2019 vs<br>2009–2010 | 2011–2013 vs<br>2009–2010 | 2014–2016 vs<br>2009–2010 | 2017–2019 vs<br>2009–2010 | 2011–2013 vs<br>2009–2010 | 2014–2016 vs<br>2009–2010 | 2017–2019 vs<br>2009–2010 |
| Males and females<br>(0–74 y) | 1.133<br>(1.081–1.188) | 1.151<br>(1.107–1.197) | 1.118<br>(1.080–1.158) | 1.053<br>(0.980–1.132) | 1.212<br>(1.136–1.295) | 1.203<br>(1.113–1.298) | 1.212<br>(1.082–1.344) | 1.312<br>(1.260–1.364) | 1.286<br>(1.236–1.338) | 1.084<br>(0.961–1.212) | 0.978<br>(0.897–1.064) | 0.956<br>(0.876–1.040) |
| Males (0–74 y) | 1.037<br>(0.937–1.143) | 1.013<br>(0.950–1.076) | 0.997<br>(0.938–1.057) | 1.067<br>(0.959–1.180) | 1.380<br>(1.311–1.449) | 1.363<br>(1.284–1.445) | 1.035<br>(0.939–1.131) | 1.099<br>(1.039–1.160) | 1.088<br>(1.043–1.133) | 1.015<br>(0.886–1.155) | 0.937<br>(0.820–1.062) | 0.948<br>(0.860–1.040) |
| Females (0–74 y) | 1.272<br>(1.210–1.337) | 1.324<br>(1.237–1.415) | 1.259<br>(1.197–1.324) | 1.151<br>(1.036–1.283) | 1.219<br>(1.094–1.368) | 1.221<br>(1.091–1.372) | 1.453<br>(1.149–1.823) | 1.577<br>(1.311–1.919) | 1.514<br>(1.272–1.824) | 1.165<br>(1.074–1.260) | 1.089<br>(1.010–1.171) | 1.046<br>(0.970–1.123) |
| Males and females<br>(0–39 y) | 1.023<br>(0.888–1.182) | 0.990<br>(0.862–1.142) | 1.137<br>(0.978–1.320) | 1.037<br>(0.846–1.270) | 1.334<br>(1.129–1.587) | 1.439<br>(1.165–1.771) | 1.285<br>(1.217–1.357) | 1.209<br>(1.107–1.313) | 1.416<br>(1.256–1.582) | 1.019<br>(0.899–1.143) | 1.000<br>(0.872–1.137) | 1.010<br>(0.899–1.126) |
| Males and females<br>(40–74 y) | 1.139<br>(1.084–1.196) | 1.160<br>(1.112–1.207) | 1.117<br>(1.078–1.157) | 1.053<br>(0.985–1.125) | 1.201<br>(1.133–1.273) | 1.182<br>(1.108–1.261) | 1.206<br>(1.068–1.343) | 1.320<br>(1.266–1.376) | 1.280<br>(1.229–1.330) | 1.137<br>(1.003–1.281) | 0.972<br>(0.902–1.047) | 0.928<br>(0.866–0.994) |
| Males (0–39 y) | 0.945<br>(0.785–1.139) | 0.799<br>(0.677–0.949) | 0.947<br>(0.794–1.136) | 0.924<br>(0.787–1.064) | 1.227<br>(1.136–1.320) | 1.232<br>(1.073–1.401) | 1.044<br>(0.839–1.321) | 1.074<br>(0.850–1.373) | 1.214<br>(0.970–1.537) | 1.050<br>(0.904–1.208) | 1.021<br>(0.851–1.202) | 1.064<br>(0.936–1.195) |
| Males (40–74 y) | 1.042<br>(0.939–1.146) | 1.026<br>(0.961–1.091) | 1.000<br>(0.939–1.061) | 1.092<br>(0.980–1.208) | 1.404<br>(1.326–1.482) | 1.385<br>(1.303–1.469) | 1.033<br>(0.933–1.136) | 1.099<br>(1.040–1.160) | 1.080<br>(1.035–1.126) | 0.998<br>(0.860–1.150) | 0.890<br>(0.796–0.997) | 0.878<br>(0.791–0.976) |
| Females (0–39 y) | 1.092<br>(0.858–1.329) | 1.195<br>(1.048–1.350) | 1.216<br>(1.099–1.337) | 1.222<br>(0.777–2.101) | 1.291<br>(0.812–2.230) | 1.505<br>(0.942–2.594) | 1.609<br>(1.131–2.345) | 1.327<br>(0.936–1.940) | 1.605<br>(1.135–2.340) | 1.009<br>(0.900–1.123) | 1.011<br>(0.895–1.135) | 0.994<br>(0.884–1.112) |
| Females (40–74 y) | 1.280<br>(1.214–1.350) | 1.330<br>(1.237–1.428) | 1.262<br>(1.200–1.327) | 1.149<br>(1.055–1.257) | 1.218<br>(1.112–1.337) | 1.209<br>(1.103–1.330) | 1.441<br>(1.139–1.806) | 1.605<br>(1.347–1.924) | 1.515<br>(1.290–1.801) | 1.283<br>(1.199–1.371) | 1.150<br>(1.069–1.233) | 1.091<br>(1.037–1.144) |

### 3.3. Age-adjusted prevalence before and after the COVID-19 outbreak

Tables 3 and S7 show the ratios and differences in age-adjusted prevalence of each disease by sex and age group among the whole of Japan before and after the COVID-19 outbreak. The prevalence of all four diseases generally increased, with the ratios among males and females aged 0–74 years being 1.058 (95% UI: 1.032–1.087) for hypertension, 1.075 (1.013–1.143) for hyperlipidemia, 1.038 (1.012–1.066) for diabetes, and 1.033 (1.027–1.039) for mental disorders. Among the age and sex groups, the ratios of actual to estimated values for these four diseases were particularly high among males aged 20–39 years and 0–39 years (ratio among males aged 0–39 years: 1.098 (1.072–1.126) for hypertension, 1.143 (1.009–1.309) for hyperlipidemia, 1.060 (1.040–1.080) for diabetes, and 1.041 (1.032–1.050) for mental disorders). In contrast, the differences in prevalence were large for males and females aged 40–64, 64–74, and 40–74 years.

**Table 3.**
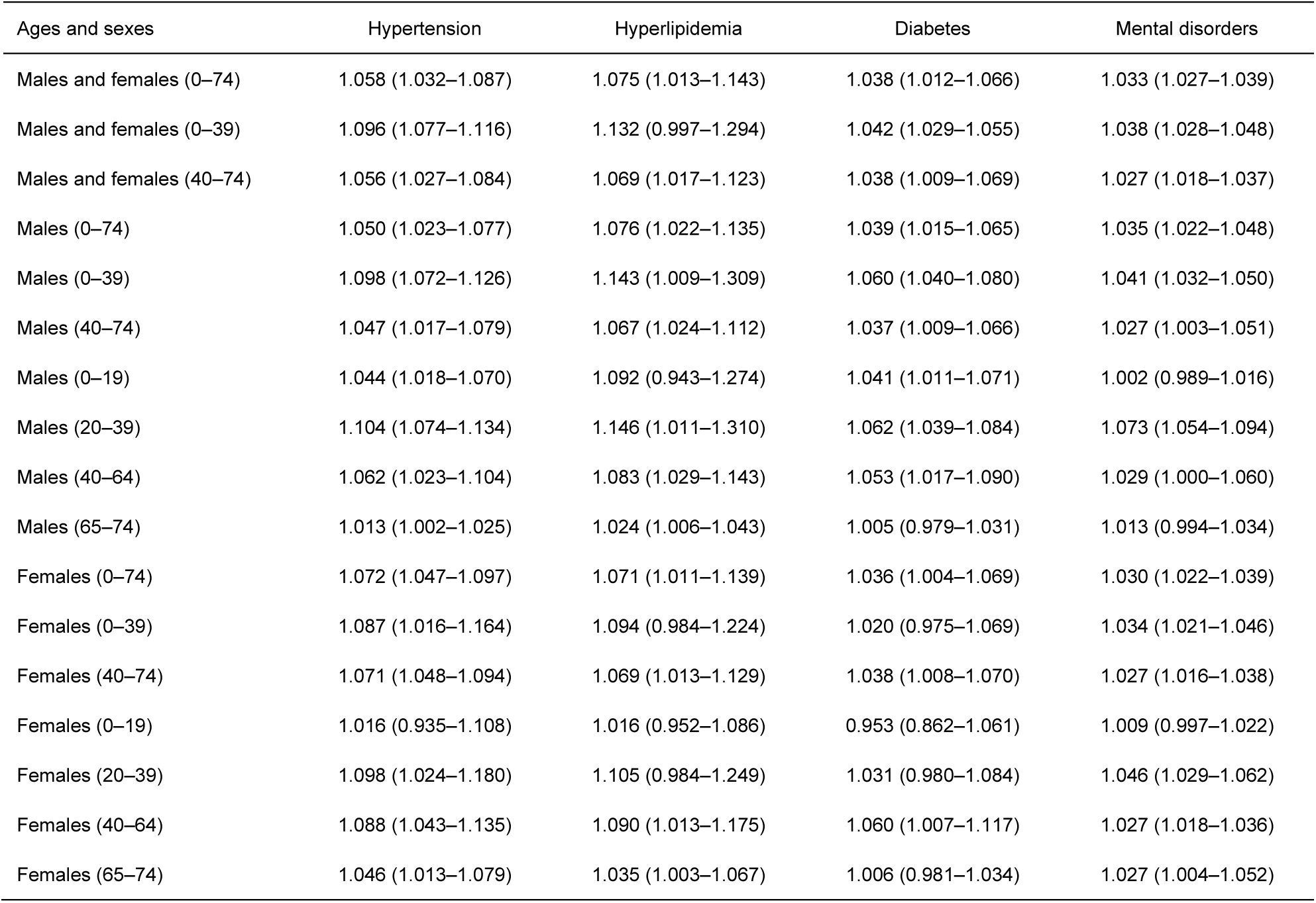
Ratio of observed to estimated prevalence in 2020 by age groups and sex (the whole of Japan). Arithmetic mean (95% uncertainty interval).

The ratios and differences in the prevalence of mental disorders before and after the COVID-19 outbreak by prefecture in males aged 0–39 years were significantly negatively correlated with the population density and COVID-19 infection rate in 2020 (Figure 4 and Tables S8 and 9). In contrast, the population density and COVID-19 infection rates were not significantly associated with the prevalence ratios and differences in other diseases.

**Figure 4.**
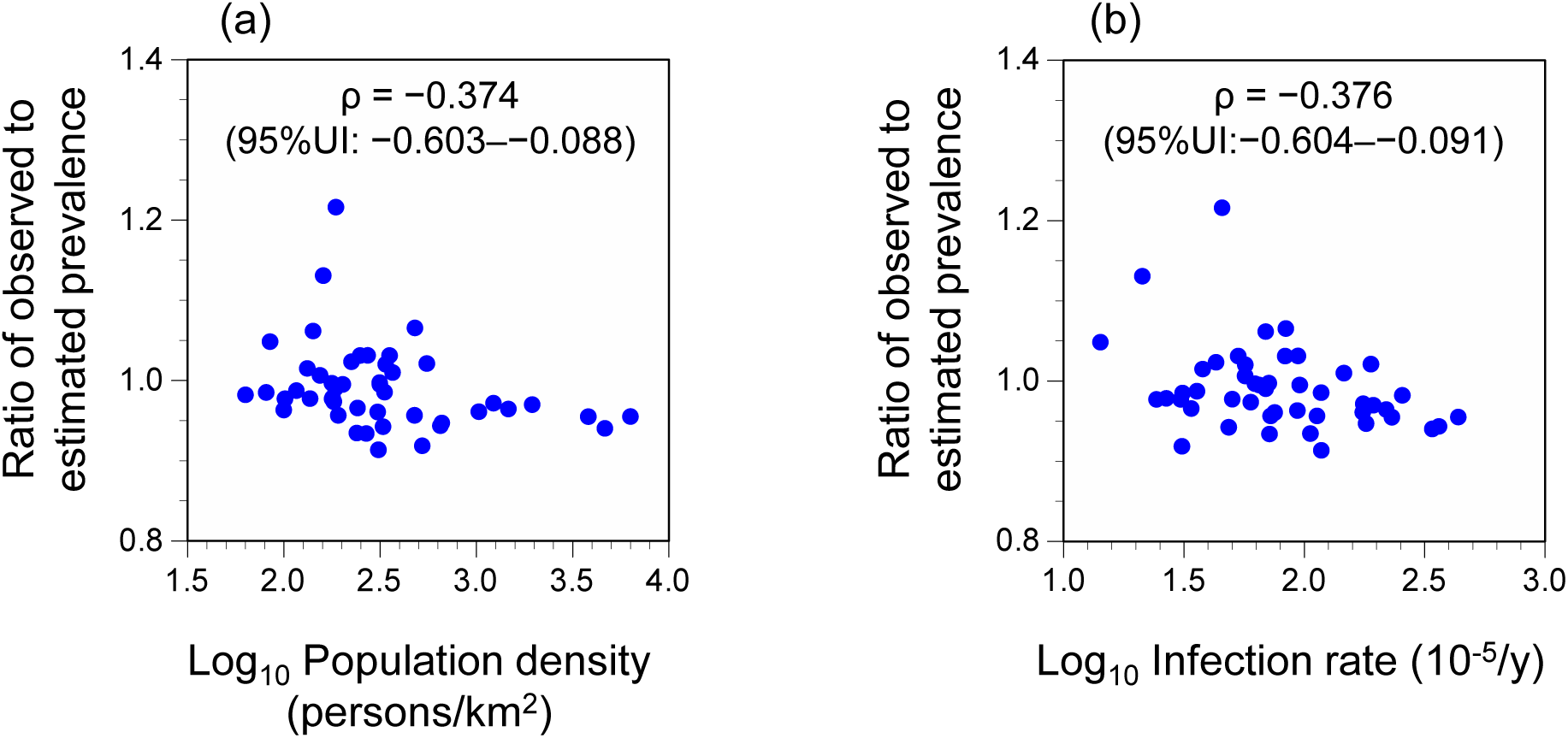
Correlations between population density or infection rate and ratio of observed to estimated age-adjusted prevalence of mental disorders in 2020 among 47 prefectures (males, 0–39 years). (a) Population density, (b) infection rate. ρ: Spearman’s rank correlation coefficient. UI: uncertainty interval.

## 4. Discussion

In this study, we evaluated the annual changes in the prevalence of hypertension, hyperlipidemia, diabetes, and mental disorders among employees of companies and their dependent family members belonging to the EHI scheme before and after the Fukushima disaster and the COVID-19 outbreak in Japan. Furthermore, we identified the vulnerable age and sex groups of people who experienced significant secondary health effects after the disaster and pandemic.

The analysis of the prevalence of these diseases after the Fukushima disaster by age and sex groups revealed that hypertension, hyperlipidemia, and diabetes generally increased in Fukushima Prefecture over the 9-year period following the disaster. Significant increases in hypertension, diabetes, and mental disorders were observed in Hamadori, hyperlipidemia and diabetes in Nakadori, and hypertension and diabetes in Aizu. Among the four diseases, the increase prevalence of diabetes was higher than that of the other diseases. Diabetes increased consistently in all the regions.

After the Fukushima disaster, a significant increase in the prevalence of diabetes was consistently reported in multiple previous studies targeting various municipalities where evacuation orders were issued [3–5, 8]. The findings of our study are consistent with those of previous studies. For hyperlipidemia, previous studies reported an increase among evacuees within 4 years after the disaster, but no significant increase in the 7-year data [3, 8]. For hypertension, some studies that used a definition based on blood pressure alone have reported a risk decrease [3, 8], whereas a study that used a definition based on blood pressure as well as the use of antihypertensive medication reported an increased risk [5]. Regarding psychological distress, a 5-fold risk increase was reported in the first year after the disaster, followed by an improvement trend [26]. Notably, this study highlighted that the worsening prevalence of these NCDs was not observed in only Hamadori, which was significantly affected by the Fukushima disaster, but also in Nakadori and Aizu. This is consistent with previous findings on non-evacuees [3, 8]. The results suggest that the Fukushima disaster may have secondary physical health effects not only on evacuees but also on the wider population. The reasons for this are not clear; however, social support for non-evacuees is more likely to be overlooked in the aftermath of a disaster than that for evacuees. Considering that these diseases generally did not change in the Iwate and Miyagi prefectures that were affected by the GJET, but not largely by the Fukushima disaster, it is possible that the increase in the prevalence of these diseases observed over a wide area in Fukushima Prefecture was related to the long-term evacuation, hosting of evacuees, and socioeconomic changes in the residents of Fukushima Prefecture following the disaster, possibly resulting in lifestyle changes among residents. Hamadori, where a significant increase in the prevalence of mental disorders was observed, includes not only municipalities whose residents were ordered to evacuate, but also those that accepted majority of evacuees. It has also been suggested that psychological distress among residents of host communities may be caused by the perception that evacuees are a threat to their social identity [9].

The increase in the prevalence of these four diseases in the post-disaster period was most pronounced among females and those aged 40–74 years. Psychological distress risk has been reported to be higher among elderly females [27]. Most of the previous studies after the Fukushima disaster have assessed the impact of evacuation on NCDs [3–5, 8], and there are limited studies that specifically discuss which age and sex groups experienced significant changes in prevalence after the disaster based on the same dataset. The strength of this study is that, based on the same dataset, changes in the prevalence after the disaster were assessed by age and sex groups to identify populations vulnerable to secondary health effects.

In contrast, after the COVID-19 outbreak, the prevalence of all four diseases increased nationwide, especially in males aged 20–39 and 0–39 years. Among the four diseases, the prevalence increase post-disaster in hyperlipidemia was particularly higher than that in the other diseases. To date, previous findings on NCDs risks after COVID-19 have not been coherent. While some population have improved their lifestyle habits [11, 12], others have decreased their physical activity levels due to lockdown and other behavioral restrictions [28]. Molarius et al. [29] reported no statistically significant changes in the prevalence of high blood pressure before and after the COVID-19 outbreak in Sweden. A systematic review regarding the impact of the COVID-19-related lockdown on glycemic control among patients with type 1 and type 2 diabetes reported that glycemic values in patients with type 1 diabetes improved, but those in patients with type 2 diabetes worsened during the COVID-19 lockdown [14]. An increasing burden of mental disorders due to the COVID-19 pandemic has been reported among the public worldwide [15]. In Japan, since the early days of the COVID-19 pandemic, active efforts have been made to raise awareness of exercise to promote physical and psychological health [30, 31]. Despite these social awareness efforts, our study showed that the prevalence of NCDs were increasing in Japan, especially among young males.

Interestingly, the age and sex groups that showed large increases in the prevalence of these diseases differed between the Fukushima disaster and COVID-19 outbreak. After the COVID-19 outbreak, not only were there risk factors such as lack of exercise and worsening of dietary and drinking habits associated with measures such as stay-at-home requests, but also preventive factors such as reduced drinking opportunities and lifestyle modification. The pandemic may have resulted in polarization, with some individuals experiencing improved health and others experiencing worsened health. The young Japanese male population tends to have lower health practices than those of other age and sex groups [32], possibly resulting in significant secondary health effects due to the pandemic. In addition, the prevalence ratios of mental disorders before and after the outbreak was significantly negatively correlated with population density and COVID-19 infection rates. This suggests that the increased prevalence in mental disorders was not mainly attributed to the burden among patients with COVID- 19, but also attributed to other social factors. Interestingly, the results of this study were in contrast to that of a previous study reporting that increases in major depressive and anxiety disorders in each country were significantly positively correlated with the rate of COVID-19 infection [15]. Although there were some differences in the timing of declaration of the state of emergency, it was issued in all prefectures in 2020 in Japan. Therefore, the differences in the impact of social activity restrictions associated with these regulations were probably small among the prefectures. There are two possible reasons for the negative association between population density and increased rate of mental disorders found in this study. First, the availability of mental health-promotion services and social infrastructure may differ among prefectures; urban areas may have had better access to mental health-promotion services and social infrastructure, even under restricted social activities during the COVID-19 pandemic. Second, there may be differences in the cultural worldviews of infection and related preventive behaviors among prefectures. The Japanese people are more likely to consider that infected individuals deserve COVID-19 infections than the people in other countries [33], which is consistent with a previous finding that Japan has a strong strict cultural view (i.e., many strong norms and low tolerance of deviant behavior) compared to that observed world [34]. It is possible that the social environment toward COVID-19 infection, shaped by this cultural worldview, had a negative impact on mental health among the population. Differences in strict cultural worldviews among prefectures in Japan and their relationship with mental disorders remain unclear and require further analysis.

This study demonstrates the importance of providing support for secondary health effects on NCDs in the aftermath of disasters and pandemics. Since significantly affected age and sex groups are unique to each disaster or pandemic, tailor-made public health support is considered important. In particular, while COVID-19 infections have a high fatality rate among the elderly [35], NCDs are more likely to affect young people. This means that infection control measures, including stay-at-home requests and other restrictions on socioeconomic activities, may be beneficial in reducing infection among the elderly under 75 years of age but may pose greater risks for the young regarding secondary health effects. Comprehensive public health measures in addition to infection control are required based on the characteristics of the population while considering the differences in the balance between risk- benefits and non-infectious secondary health effects among the populations.

This study has some limitations. First, the target population was employees of companies and their dependent family members. The findings of this study cannot be generalized to the entire population and other countries. Furthermore, only the population aged 74 years and below were included in this study. Although COVID-19 isolation measures are known to worsen cognitive and mental health among people with dementia [36], this study was unable to assess the secondary health effects among the population aged 75 years and older. Second, we could not fully examine the possibility of changes in accessibility to healthcare facilities after the Fukushima disaster or the COVID-19 outbreak compared to accessibility to health care before the disaster and outbreak. However, it is unlikely that NCDs were overlooked before the disaster or outbreak, since employees of companies, the target population of this study, generally receive regular health checkups. Third, this study was based on an ecological study design and did not identify the causes of changes in the prevalence of various diseases. Long-term cohort studies based on individuals exposed to disasters and pandemics are expected to assess the impact of disasters and pandemics on secondary health and identify the factors that influence this impact.

Despite these limitations, this study provided unique insights into the secondary health effects of the Fukushima disaster and COVID-19 outbreak and identified age and sex groups that were significantly affected by this disaster and pandemic.

## 5. Conclusions

This study used health insurance receipt data for employees of companies and their dependent family members from January 2009 to December 2020 to assess changes in the prevalence of NCDs after the Fukushima disaster and COVID-19 outbreak in Japan compared to that before the disaster and outbreak and to identify age and sex groups particularly vulnerable to the resultant secondary health effects. The findings of this study are summarized as follows.

- Age-adjusted prevalence of hypertension, hyperlipidemia, and diabetes increased over a 9-year period in the Fukushima Prefecture following the 2011 Fukushima disaster. An increase in the prevalence in mental disorders was also observed in the Hamadori region in the prefecture, which was most affected by the disaster.
- Increase prevalence in hypertension, hyperlipidemia, and diabetes were also observed in Hamadori and Aizu, which were less affected by the disaster, indicating that the Fukushima disaster had secondary health effects over wider populations.
- In contrast, no significant changes were observed for these four diseases in Iwate and Miyagi prefectures.
- After the Fukushima disaster, the prevalence ratios of these four diseases were particularly high among females aged 40–74 years.
- After the COVID-19 outbreak, the prevalence of these four diseases has increased significantly throughout Japan. In particular, the post-outbreak prevalence ratios of these four diseases were significantly higher among males aged 0–39 years than among females and males of other age groups, which differed from the age and sex groups vulnerable to secondary health effects after the Fukushima disaster.
- Post-outbreak prevalence ratios of mental disorders among males aged 0–39 years by prefecture were negatively and significantly associated with population density and COVID-19 infection rates.
- The population groups vulnerable to secondary health effects on NCDs after the Fukushima disaster or the COVID-19 outbreak are not common but are unique to the disaster or pandemic. It is important to provide tailor-made public health support to the population accordance to disasters and pandemics.

## Data Availability

The raw data used in the study were provided by JMDC Inc., as described in this paper. We are unable to attach all of the raw data in this paper, because it would violate the contract; however, we have included the crude prevalence data needed to replicate the study and all data produced in the present work in the manuscript and Supplementary materials.

## Acknowledgements

This work was supported by JSPS KAKENHI grant number JP20H04354. We would like to thank Editage (www.editage.com) for English language editing and Dr. Kazuki Ide (Osaka University) for his advice.

## Authorship contribution

Michio Murakami: Conceptualization; Data curation; Methodology; Formal analysis; Visualization; Project administration; Writing –original draft Shuhei Nomura: Conceptualization; Writing –review & editing

## Supplementary Materials

**Table S1.** The number of diseases from 2009 to 2020 (the whole of Japan, males, and females; 0–74 years).

| Area | Sex | Ages | Diseases | 2009 | 2010 | 2011 | 2012 | 2013 | 2014 | 2015 | 2016 | 2017 | 2018 | 2019 | 2020 |
| --- | --- | --- | --- | --- | --- | --- | --- | --- | --- | --- | --- | --- | --- | --- | --- |
| The whole of Japan | Males and females | 0–74 | Hypertension | 35223 | 48281 | 79751 | 127912 | 157304 | 234540 | 247773 | 391114 | 525666 | 626767 | 714748 | 781664 |
| The whole of Japan | Males and females | 0–74 | Hyperlipidemia | 39005 | 52673 | 88783 | 142425 | 173314 | 259782 | 277760 | 425027 | 559295 | 677761 | 786923 | 853000 |
| The whole of Japan | Males and females | 0–74 | Diabetes | 21960 | 29492 | 48317 | 75415 | 91201 | 137408 | 147285 | 233631 | 314817 | 379289 | 434864 | 468609 |
| The whole of Japan | Males and females | 0–74 | Mental disorders | 32207 | 44208 | 75891 | 112707 | 133637 | 203871 | 220154 | 334829 | 456795 | 564433 | 643366 | 677042 |
| The whole of Japan | Males and females | 0–74 | Any types of diseases | 462365 | 575117 | 989516 | 1411998 | 1641456 | 2453223 | 2589088 | 3791740 | 5065073 | 6094545 | 6815288 | 6786523 |

**Table S2.** Annual age-adjusted prevalence of each disease in Iwate, Miyagi, Fukushima prefectures, Hamadori, Nakadori, Aizu, and the whole of Japan.

| Area | Sex | Ages | Diseases | 2009 | 2010 | 2011 | 2012 | 2013 | 2014 | 2015 | 2016 | 2017 | 2018 | 2019 | 2020 |
| --- | --- | --- | --- | --- | --- | --- | --- | --- | --- | --- | --- | --- | --- | --- | --- |
| The whole of Japan | Males and females | 0–74 | Hypertension | 0.102 | 0.106 | 0.103 | 0.105 | 0.107 | 0.106 | 0.104 | 0.106 | 0.106 | 0.105 | 0.107 | 0.113 |
| Iwate Prefecture | Males and females | 0–74 | Hypertension | 0.119 | 0.126 | 0.125 | 0.134 | 0.129 | 0.117 | 0.116 | 0.115 | 0.114 | 0.116 | 0.120 | 0.120 |
| Miyagi Prefecture | Males and females | 0–74 | Hypertension | 0.128 | 0.117 | 0.114 | 0.126 | 0.124 | 0.122 | 0.120 | 0.113 | 0.111 | 0.114 | 0.115 | 0.113 |
| Fukushima Prefecture | Males and females | 0–74 | Hypertension | 0.106 | 0.104 | 0.123 | 0.120 | 0.117 | 0.125 | 0.122 | 0.121 | 0.119 | 0.119 | 0.121 | 0.117 |
| Hamadori | Males and females | 0–74 | Hypertension | 0.131 | 0.130 | 0.179 | 0.152 | 0.156 | 0.144 | 0.148 | 0.144 | 0.144 | 0.151 | 0.150 | 0.142 |
| Nakadori | Males and females | 0–74 | Hypertension | 0.133 | 0.120 | 0.120 | 0.122 | 0.122 | 0.135 | 0.134 | 0.134 | 0.128 | 0.126 | 0.129 | 0.124 |
| Aizu | Males and females | 0–74 | Hypertension | 0.112 | 0.107 | 0.117 | 0.124 | 0.131 | 0.124 | 0.122 | 0.122 | 0.121 | 0.119 | 0.116 | 0.116 |
| The whole of Japan | Males and females | 0–74 | Hyperlipidemia | 0.107 | 0.110 | 0.109 | 0.112 | 0.114 | 0.114 | 0.114 | 0.113 | 0.111 | 0.112 | 0.115 | 0.121 |
| Iwate Prefecture | Males and females | 0–74 | Hyperlipidemia | 0.100 | 0.123 | 0.125 | 0.133 | 0.136 | 0.130 | 0.131 | 0.132 | 0.128 | 0.131 | 0.135 | 0.131 |
| Miyagi Prefecture | Males and females | 0–74 | Hyperlipidemia | 0.115 | 0.103 | 0.115 | 0.125 | 0.127 | 0.120 | 0.119 | 0.117 | 0.115 | 0.120 | 0.121 | 0.117 |
| Fukushima Prefecture | Males and females | 0–74 | Hyperlipidemia | 0.098 | 0.092 | 0.104 | 0.102 | 0.103 | 0.120 | 0.121 | 0.120 | 0.116 | 0.116 | 0.122 | 0.119 |
| Hamadori | Males and females | 0–74 | Hyperlipidemia | 0.153 | 0.128 | 0.134 | 0.155 | 0.146 | 0.162 | 0.160 | 0.150 | 0.147 | 0.150 | 0.154 | 0.150 |
| Nakadori | Males and females | 0–74 | Hyperlipidemia | 0.106 | 0.091 | 0.096 | 0.103 | 0.110 | 0.135 | 0.137 | 0.136 | 0.129 | 0.128 | 0.135 | 0.130 |
| Aizu | Males and females | 0–74 | Hyperlipidemia | 0.121 | 0.116 | 0.112 | 0.109 | 0.116 | 0.114 | 0.114 | 0.123 | 0.126 | 0.119 | 0.126 | 0.118 |
| The whole of Japan | Males and females | 0–74 | Diabetes | 0.061 | 0.063 | 0.061 | 0.061 | 0.062 | 0.062 | 0.062 | 0.064 | 0.064 | 0.064 | 0.065 | 0.068 |
| Iwate Prefecture | Males and females | 0–74 | Diabetes | 0.059 | 0.055 | 0.066 | 0.065 | 0.064 | 0.055 | 0.056 | 0.059 | 0.060 | 0.060 | 0.060 | 0.060 |
| Miyagi Prefecture | Males and females | 0–74 | Diabetes | 0.076 | 0.059 | 0.066 | 0.071 | 0.076 | 0.070 | 0.070 | 0.067 | 0.066 | 0.068 | 0.068 | 0.065 |
| Fukushima Prefecture | Males and females | 0–74 | Diabetes | 0.044 | 0.043 | 0.047 | 0.052 | 0.056 | 0.057 | 0.057 | 0.057 | 0.057 | 0.057 | 0.058 | 0.056 |
| Hamadori | Males and females | 0–74 | Diabetes | 0.051 | 0.067 | 0.065 | 0.080 | 0.081 | 0.080 | 0.078 | 0.075 | 0.072 | 0.074 | 0.070 | 0.070 |
| Nakadori | Males and females | 0–74 | Diabetes | 0.047 | 0.043 | 0.042 | 0.047 | 0.058 | 0.059 | 0.063 | 0.061 | 0.062 | 0.062 | 0.064 | 0.060 |
| Aizu | Males and females | 0–74 | Diabetes | 0.049 | 0.048 | 0.044 | 0.051 | 0.054 | 0.055 | 0.054 | 0.066 | 0.062 | 0.060 | 0.063 | 0.064 |
| The whole of Japan | Males and females | 0–74 | Mental disorders | 0.075 | 0.081 | 0.080 | 0.082 | 0.083 | 0.084 | 0.086 | 0.088 | 0.089 | 0.091 | 0.093 | 0.098 |
| Iwate Prefecture | Males and females | 0–74 | Mental disorders | 0.057 | 0.086 | 0.077 | 0.068 | 0.069 | 0.067 | 0.068 | 0.071 | 0.072 | 0.077 | 0.077 | 0.078 |
| Miyagi Prefecture | Males and females | 0–74 | Mental disorders | 0.073 | 0.084 | 0.075 | 0.077 | 0.081 | 0.077 | 0.078 | 0.081 | 0.080 | 0.081 | 0.081 | 0.081 |
| Fukushima Prefecture | Males and females | 0–74 | Mental disorders | 0.072 | 0.075 | 0.090 | 0.081 | 0.078 | 0.077 | 0.078 | 0.080 | 0.081 | 0.083 | 0.082 | 0.082 |
| Hamadori | Males and females | 0–74 | Mental disorders | 0.064 | 0.069 | 0.114 | 0.083 | 0.078 | 0.097 | 0.085 | 0.094 | 0.095 | 0.110 | 0.103 | 0.101 |
| Nakadori | Males and females | 0–74 | Mental disorders | 0.088 | 0.086 | 0.097 | 0.096 | 0.091 | 0.087 | 0.087 | 0.097 | 0.098 | 0.096 | 0.096 | 0.095 |
| Aizu | Males and females | 0–74 | Mental disorders | 0.081 | 0.085 | 0.094 | 0.087 | 0.091 | 0.091 | 0.094 | 0.092 | 0.097 | 0.092 | 0.094 | 0.087 |
| The whole of Japan | Males and females | 0–39 | Hypertension | 0.010 | 0.010 | 0.009 | 0.010 | 0.010 | 0.010 | 0.010 | 0.010 | 0.010 | 0.010 | 0.010 | 0.011 |
| Fukushima Prefecture | Males and females | 0–39 | Hypertension | 0.010 | 0.008 | 0.009 | 0.009 | 0.009 | 0.009 | 0.008 | 0.009 | 0.009 | 0.010 | 0.011 | 0.010 |
| The whole of Japan | Males and females | 0–39 | Hyperlipidemia | 0.019 | 0.018 | 0.018 | 0.019 | 0.019 | 0.019 | 0.020 | 0.019 | 0.019 | 0.019 | 0.021 | 0.023 |
| Fukushima Prefecture | Males and females | 0–39 | Hyperlipidemia | 0.014 | 0.012 | 0.012 | 0.015 | 0.014 | 0.018 | 0.017 | 0.019 | 0.018 | 0.019 | 0.022 | 0.022 |
| The whole of Japan | Males and females | 0–39 | Diabetes | 0.009 | 0.009 | 0.009 | 0.009 | 0.009 | 0.009 | 0.009 | 0.010 | 0.010 | 0.011 | 0.011 | 0.012 |
| Fukushima Prefecture | Males and females | 0–39 | Diabetes | 0.005 | 0.005 | 0.006 | 0.006 | 0.006 | 0.006 | 0.005 | 0.006 | 0.007 | 0.008 | 0.008 | 0.009 |
| The whole of Japan | Males and females | 0–39 | Mental disorders | 0.054 | 0.060 | 0.061 | 0.063 | 0.064 | 0.066 | 0.068 | 0.071 | 0.073 | 0.077 | 0.080 | 0.086 |
| Fukushima Prefecture | Males and females | 0–39 | Mental disorders | 0.048 | 0.049 | 0.057 | 0.052 | 0.053 | 0.054 | 0.058 | 0.062 | 0.064 | 0.067 | 0.066 | 0.068 |
| The whole of Japan | Males and females | 40–74 | Hypertension | 0.244 | 0.252 | 0.246 | 0.250 | 0.255 | 0.253 | 0.249 | 0.254 | 0.252 | 0.251 | 0.254 | 0.268 |
| Fukushima Prefecture | Males and females | 40–74 | Hypertension | 0.253 | 0.251 | 0.297 | 0.289 | 0.282 | 0.302 | 0.296 | 0.292 | 0.286 | 0.286 | 0.288 | 0.282 |
| The whole of Japan | Males and females | 40–74 | Hyperlipidemia | 0.242 | 0.249 | 0.247 | 0.255 | 0.259 | 0.258 | 0.258 | 0.256 | 0.251 | 0.252 | 0.259 | 0.272 |
| Fukushima Prefecture | Males and females | 40–74 | Hyperlipidemia | 0.225 | 0.214 | 0.244 | 0.235 | 0.238 | 0.277 | 0.278 | 0.273 | 0.265 | 0.265 | 0.275 | 0.268 |
| The whole of Japan | Males and females | 40–74 | Diabetes | 0.141 | 0.145 | 0.141 | 0.141 | 0.142 | 0.142 | 0.142 | 0.146 | 0.145 | 0.145 | 0.147 | 0.154 |
| Fukushima Prefecture | Males and females | 40–74 | Diabetes | 0.103 | 0.101 | 0.110 | 0.121 | 0.133 | 0.136 | 0.135 | 0.134 | 0.132 | 0.133 | 0.134 | 0.129 |
| The whole of Japan | Males and females | 40–74 | Mental disorders | 0.107 | 0.112 | 0.110 | 0.111 | 0.110 | 0.111 | 0.113 | 0.113 | 0.113 | 0.113 | 0.113 | 0.117 |
| Fukushima Prefecture | Males and females | 40–74 | Mental disorders | 0.108 | 0.113 | 0.140 | 0.126 | 0.117 | 0.114 | 0.109 | 0.108 | 0.106 | 0.107 | 0.106 | 0.103 |
| The whole of Japan | Males | 0–74 | Hypertension | 0.114 | 0.120 | 0.117 | 0.120 | 0.124 | 0.125 | 0.124 | 0.128 | 0.128 | 0.129 | 0.131 | 0.139 |
| Fukushima Prefecture | Males | 0–74 | Hypertension | 0.126 | 0.122 | 0.142 | 0.129 | 0.127 | 0.136 | 0.134 | 0.135 | 0.136 | 0.137 | 0.138 | 0.135 |
| The whole of Japan | Males | 0-74 | Hyperlipidemia | 0.109 | 0.113 | 0.114 | 0.121 | 0.125 | 0.125 | 0.126 | 0.126 | 0.125 | 0.127 | 0.131 | 0.140 |
| Fukushima Prefecture | Males | 0-74 | Hyperlipidemia | 0.081 | 0.081 | 0.086 | 0.096 | 0.099 | 0.123 | 0.127 | 0.129 | 0.125 | 0.125 | 0.131 | 0.129 |
| The whole of Japan | Males | 0-74 | Diabetes | 0.073 | 0.076 | 0.072 | 0.072 | 0.073 | 0.073 | 0.074 | 0.077 | 0.077 | 0.078 | 0.079 | 0.084 |
| Fukushima Prefecture | Males | 0-74 | Diabetes | 0.059 | 0.059 | 0.055 | 0.059 | 0.064 | 0.063 | 0.066 | 0.066 | 0.067 | 0.067 | 0.067 | 0.065 |
| The whole of Japan | Males | 0-74 | Mental disorders | 0.071 | 0.076 | 0.077 | 0.079 | 0.079 | 0.081 | 0.084 | 0.086 | 0.088 | 0.090 | 0.091 | 0.097 |
| Fukushima Prefecture | Males | 0-74 | Mental disorders | 0.063 | 0.066 | 0.077 | 0.066 | 0.067 | 0.063 | 0.071 | 0.073 | 0.074 | 0.076 | 0.074 | 0.074 |
| The whole of Japan | Males | 0-39 | Hypertension | 0.013 | 0.013 | 0.012 | 0.012 | 0.012 | 0.012 | 0.012 | 0.012 | 0.012 | 0.013 | 0.013 | 0.014 |
| Fukushima Prefecture | Males | 0-39 | Hypertension | 0.014 | 0.012 | 0.013 | 0.012 | 0.011 | 0.010 | 0.010 | 0.010 | 0.011 | 0.012 | 0.013 | 0.012 |
| The whole of Japan | Males | 0-39 | Hyperlipidemia | 0.024 | 0.024 | 0.022 | 0.024 | 0.024 | 0.024 | 0.025 | 0.024 | 0.024 | 0.025 | 0.027 | 0.030 |
| Fukushima Prefecture | Males | 0-39 | Hyperlipidemia | 0.019 | 0.019 | 0.015 | 0.019 | 0.019 | 0.023 | 0.023 | 0.026 | 0.023 | 0.024 | 0.027 | 0.028 |
| The whole of Japan | Males | 0-39 | Diabetes | 0.011 | 0.011 | 0.010 | 0.010 | 0.010 | 0.010 | 0.010 | 0.011 | 0.011 | 0.012 | 0.012 | 0.013 |
| Fukushima Prefecture | Males | 0-39 | Diabetes | 0.006 | 0.007 | 0.006 | 0.005 | 0.006 | 0.006 | 0.006 | 0.007 | 0.007 | 0.008 | 0.009 | 0.009 |
| The whole of Japan | Males | 0-39 | Mental disorders | 0.056 | 0.062 | 0.064 | 0.066 | 0.068 | 0.070 | 0.073 | 0.076 | 0.078 | 0.082 | 0.084 | 0.091 |
| Fukushima Prefecture | Males | 0-39 | Mental disorders | 0.045 | 0.044 | 0.056 | 0.049 | 0.051 | 0.049 | 0.057 | 0.061 | 0.063 | 0.067 | 0.065 | 0.067 |
| The whole of Japan | Males | 40-74 | Hypertension | 0.267 | 0.283 | 0.277 | 0.284 | 0.295 | 0.296 | 0.295 | 0.304 | 0.305 | 0.306 | 0.311 | 0.329 |
| Fukushima Prefecture | Males | 40-74 | Hypertension | 0.297 | 0.290 | 0.339 | 0.307 | 0.304 | 0.328 | 0.324 | 0.327 | 0.327 | 0.328 | 0.329 | 0.323 |
| The whole of Japan | Males | 40-74 | Hyperlipidemia | 0.238 | 0.251 | 0.255 | 0.270 | 0.278 | 0.278 | 0.281 | 0.281 | 0.279 | 0.283 | 0.290 | 0.308 |
| Fukushima Prefecture | Males | 40-74 | Hyperlipidemia | 0.176 | 0.176 | 0.195 | 0.213 | 0.221 | 0.277 | 0.286 | 0.286 | 0.282 | 0.279 | 0.289 | 0.282 |
| The whole of Japan | Males | 40-74 | Diabetes | 0.168 | 0.176 | 0.168 | 0.168 | 0.169 | 0.171 | 0.172 | 0.177 | 0.178 | 0.179 | 0.181 | 0.190 |
| Fukushima Prefecture | Males | 40-74 | Diabetes | 0.140 | 0.140 | 0.131 | 0.140 | 0.153 | 0.150 | 0.157 | 0.157 | 0.158 | 0.157 | 0.157 | 0.151 |
| The whole of Japan | Males | 40-74 | Mental disorders | 0.093 | 0.098 | 0.097 | 0.099 | 0.097 | 0.098 | 0.100 | 0.102 | 0.102 | 0.102 | 0.102 | 0.105 |
| Fukushima Prefecture | Males | 40-74 | Mental disorders | 0.091 | 0.100 | 0.108 | 0.094 | 0.091 | 0.085 | 0.091 | 0.091 | 0.092 | 0.090 | 0.087 | 0.085 |
| The whole of Japan | Females | 0-74 | Hypertension | 0.087 | 0.089 | 0.086 | 0.086 | 0.086 | 0.084 | 0.082 | 0.082 | 0.081 | 0.079 | 0.080 | 0.084 |
| Fukushima Prefecture | Females | 0-74 | Hypertension | 0.088 | 0.084 | 0.107 | 0.108 | 0.106 | 0.111 | 0.108 | 0.103 | 0.098 | 0.099 | 0.099 | 0.096 |
| The whole of Japan | Females | 0–74 | Hyperlipidemia | 0.101 | 0.103 | 0.100 | 0.101 | 0.101 | 0.100 | 0.099 | 0.099 | 0.095 | 0.095 | 0.097 | 0.102 |
| Fukushima Prefecture | Females | 0–74 | Hyperlipidemia | 0.097 | 0.087 | 0.105 | 0.104 | 0.103 | 0.112 | 0.108 | 0.106 | 0.103 | 0.104 | 0.109 | 0.107 |
| The whole of Japan | Females | 0–74 | Diabetes | 0.048 | 0.049 | 0.048 | 0.048 | 0.048 | 0.048 | 0.048 | 0.049 | 0.048 | 0.049 | 0.049 | 0.051 |
| Fukushima Prefecture | Females | 0–74 | Diabetes | 0.033 | 0.027 | 0.037 | 0.044 | 0.049 | 0.050 | 0.046 | 0.045 | 0.044 | 0.046 | 0.047 | 0.046 |
| The whole of Japan | Females | 0–74 | Mental disorders | 0.080 | 0.084 | 0.084 | 0.085 | 0.087 | 0.087 | 0.088 | 0.090 | 0.091 | 0.093 | 0.095 | 0.100 |
| Fukushima Prefecture | Females | 0–74 | Mental disorders | 0.074 | 0.076 | 0.093 | 0.093 | 0.087 | 0.091 | 0.086 | 0.089 | 0.087 | 0.091 | 0.091 | 0.091 |
| The whole of Japan | Females | 0–39 | Hypertension | 0.006 | 0.006 | 0.006 | 0.007 | 0.007 | 0.007 | 0.007 | 0.007 | 0.007 | 0.007 | 0.007 | 0.008 |
| Fukushima Prefecture | Females | 0–39 | Hypertension | 0.006 | 0.006 | 0.006 | 0.006 | 0.008 | 0.007 | 0.007 | 0.009 | 0.007 | 0.008 | 0.008 | 0.007 |
| The whole of Japan | Females | 0–39 | Hyperlipidemia | 0.013 | 0.013 | 0.012 | 0.013 | 0.013 | 0.013 | 0.013 | 0.014 | 0.013 | 0.013 | 0.014 | 0.015 |
| Fukushima Prefecture | Females | 0–39 | Hyperlipidemia | 0.011 | 0.007 | 0.010 | 0.011 | 0.009 | 0.013 | 0.011 | 0.010 | 0.012 | 0.013 | 0.015 | 0.015 |
| The whole of Japan | Females | 0–39 | Diabetes | 0.008 | 0.008 | 0.008 | 0.008 | 0.008 | 0.008 | 0.009 | 0.009 | 0.009 | 0.009 | 0.010 | 0.010 |
| Fukushima Prefecture | Females | 0–39 | Diabetes | 0.004 | 0.003 | 0.005 | 0.007 | 0.006 | 0.006 | 0.005 | 0.005 | 0.006 | 0.007 | 0.008 | 0.009 |
| The whole of Japan | Females | 0–39 | Mental disorders | 0.053 | 0.057 | 0.057 | 0.059 | 0.061 | 0.062 | 0.063 | 0.066 | 0.068 | 0.072 | 0.075 | 0.080 |
| Fukushima Prefecture | Females | 0–39 | Mental disorders | 0.051 | 0.054 | 0.058 | 0.056 | 0.055 | 0.059 | 0.059 | 0.063 | 0.066 | 0.069 | 0.067 | 0.069 |
| The whole of Japan | Females | 40–74 | Hypertension | 0.210 | 0.215 | 0.208 | 0.208 | 0.206 | 0.202 | 0.196 | 0.196 | 0.193 | 0.190 | 0.191 | 0.202 |
| Fukushima Prefecture | Females | 40–74 | Hypertension | 0.213 | 0.204 | 0.262 | 0.265 | 0.256 | 0.269 | 0.261 | 0.247 | 0.237 | 0.237 | 0.238 | 0.232 |
| The whole of Japan | Females | 40–74 | Hyperlipidemia | 0.236 | 0.240 | 0.234 | 0.235 | 0.235 | 0.233 | 0.229 | 0.229 | 0.221 | 0.220 | 0.224 | 0.234 |
| Fukushima Prefecture | Females | 40–74 | Hyperlipidemia | 0.228 | 0.209 | 0.250 | 0.245 | 0.246 | 0.263 | 0.257 | 0.251 | 0.241 | 0.242 | 0.253 | 0.248 |
| The whole of Japan | Females | 40–74 | Diabetes | 0.109 | 0.112 | 0.110 | 0.109 | 0.110 | 0.109 | 0.108 | 0.110 | 0.108 | 0.108 | 0.110 | 0.113 |
| Fukushima Prefecture | Females | 40–74 | Diabetes | 0.076 | 0.065 | 0.086 | 0.101 | 0.114 | 0.117 | 0.109 | 0.105 | 0.102 | 0.105 | 0.106 | 0.102 |
| The whole of Japan | Females | 40–74 | Mental disorders | 0.120 | 0.125 | 0.123 | 0.126 | 0.126 | 0.126 | 0.127 | 0.127 | 0.126 | 0.126 | 0.127 | 0.130 |
| Fukushima Prefecture | Females | 40–74 | Mental disorders | 0.110 | 0.111 | 0.146 | 0.150 | 0.138 | 0.139 | 0.127 | 0.127 | 0.121 | 0.126 | 0.127 | 0.123 |

**Table S3.** Annual age-adjusted prevalence of each disease in the whole of Japan by age groups and sex and 47 prefectures (males, 0–39 years) from 2015 to 2020.

| Area | Sex | Ages | Diseases | 2015 | 2016 | 2017 | 2018 | 2019 | 2020 |
| --- | --- | --- | --- | --- | --- | --- | --- | --- | --- |
| The whole of Japan | Males and females | 0–39 | Hypertension | 0.010 | 0.010 | 0.010 | 0.010 | 0.010 | 0.011 |
| The whole of Japan | Males and females | 40–74 | Hypertension | 0.249 | 0.254 | 0.252 | 0.251 | 0.254 | 0.268 |
| The whole of Japan | Males and females | 0–74 | Hypertension | 0.104 | 0.106 | 0.106 | 0.105 | 0.107 | 0.113 |
| The whole of Japan | Males | 0–39 | Hypertension | 0.012 | 0.012 | 0.012 | 0.013 | 0.013 | 0.014 |
| The whole of Japan | Males | 40–74 | Hypertension | 0.295 | 0.304 | 0.305 | 0.306 | 0.311 | 0.329 |
| The whole of Japan | Males | 0–74 | Hypertension | 0.124 | 0.128 | 0.128 | 0.129 | 0.131 | 0.139 |
| The whole of Japan | Males | 0–19 | Hypertension | 0.002 | 0.002 | 0.002 | 0.002 | 0.002 | 0.003 |
| The whole of Japan | Males | 20–39 | Hypertension | 0.021 | 0.021 | 0.022 | 0.022 | 0.022 | 0.025 |
| The whole of Japan | Males | 40–64 | Hypertension | 0.245 | 0.254 | 0.255 | 0.255 | 0.259 | 0.278 |
| The whole of Japan | Males | 65–74 | Hypertension | 0.529 | 0.539 | 0.545 | 0.548 | 0.555 | 0.568 |
| The whole of Japan | Females | 0–39 | Hypertension | 0.007 | 0.007 | 0.007 | 0.007 | 0.007 | 0.008 |
| The whole of Japan | Females | 40–74 | Hypertension | 0.196 | 0.196 | 0.193 | 0.190 | 0.191 | 0.202 |
| The whole of Japan | Females | 0–74 | Hypertension | 0.082 | 0.082 | 0.081 | 0.079 | 0.080 | 0.084 |
| The whole of Japan | Females | 0–19 | Hypertension | 0.001 | 0.002 | 0.002 | 0.002 | 0.002 | 0.002 |
| The whole of Japan | Females | 20–39 | Hypertension | 0.012 | 0.012 | 0.012 | 0.012 | 0.012 | 0.013 |
| The whole of Japan | Females | 40–64 | Hypertension | 0.136 | 0.139 | 0.137 | 0.135 | 0.137 | 0.148 |
| The whole of Japan | Females | 65–74 | Hypertension | 0.481 | 0.466 | 0.457 | 0.452 | 0.448 | 0.457 |
| The whole of Japan | Males and females | 0–39 | Hyperlipidemia | 0.020 | 0.019 | 0.019 | 0.019 | 0.021 | 0.023 |
| The whole of Japan | Males and females | 40–74 | Hyperlipidemia | 0.258 | 0.256 | 0.251 | 0.252 | 0.259 | 0.272 |
| The whole of Japan | Males and females | 0–74 | Hyperlipidemia | 0.114 | 0.113 | 0.111 | 0.112 | 0.115 | 0.121 |
| The whole of Japan | Males | 0–39 | Hyperlipidemia | 0.025 | 0.024 | 0.024 | 0.025 | 0.027 | 0.030 |
| The whole of Japan | Males | 40–74 | Hyperlipidemia | 0.281 | 0.281 | 0.279 | 0.283 | 0.290 | 0.308 |
| The whole of Japan | Males | 0–74 | Hyperlipidemia | 0.126 | 0.126 | 0.125 | 0.127 | 0.131 | 0.140 |
| The whole of Japan | Males | 0–19 | Hyperlipidemia | 0.003 | 0.003 | 0.003 | 0.003 | 0.003 | 0.004 |
| The whole of Japan | Males | 20–39 | Hyperlipidemia | 0.045 | 0.044 | 0.043 | 0.046 | 0.049 | 0.055 |
| The whole of Japan | Males | 40–64 | Hyperlipidemia | 0.252 | 0.252 | 0.247 | 0.251 | 0.258 | 0.277 |
| The whole of Japan | Males | 65–74 | Hyperlipidemia | 0.420 | 0.421 | 0.427 | 0.434 | 0.441 | 0.456 |
| The whole of Japan | Females | 0–39 | Hyperlipidemia | 0.013 | 0.014 | 0.013 | 0.013 | 0.014 | 0.015 |
| The whole of Japan | Females | 40–74 | Hyperlipidemia | 0.229 | 0.229 | 0.221 | 0.220 | 0.224 | 0.234 |
| The whole of Japan | Females | 0–74 | Hyperlipidemia | 0.099 | 0.099 | 0.095 | 0.095 | 0.097 | 0.102 |
| The whole of Japan | Females | 0–19 | Hyperlipidemia | 0.003 | 0.003 | 0.003 | 0.003 | 0.003 | 0.003 |
| The whole of Japan | Females | 20–39 | Hyperlipidemia | 0.023 | 0.023 | 0.022 | 0.023 | 0.025 | 0.027 |
| The whole of Japan | Females | 40–64 | Hyperlipidemia | 0.171 | 0.172 | 0.165 | 0.164 | 0.169 | 0.179 |
| The whole of Japan | Females | 65–74 | Hyperlipidemia | 0.506 | 0.496 | 0.486 | 0.484 | 0.484 | 0.491 |
| The whole of Japan | Males and females | 0–39 | Diabetes | 0.009 | 0.010 | 0.010 | 0.011 | 0.011 | 0.012 |
| The whole of Japan | Males and females | 40–74 | Diabetes | 0.142 | 0.146 | 0.145 | 0.145 | 0.147 | 0.154 |
| The whole of Japan | Males and females | 0–74 | Diabetes | 0.062 | 0.064 | 0.064 | 0.064 | 0.065 | 0.068 |
| The whole of Japan | Males | 0–39 | Diabetes | 0.010 | 0.011 | 0.011 | 0.012 | 0.012 | 0.013 |
| The whole of Japan | Males | 40–74 | Diabetes | 0.172 | 0.177 | 0.178 | 0.179 | 0.181 | 0.190 |
| The whole of Japan | Males | 0–74 | Diabetes | 0.074 | 0.077 | 0.077 | 0.078 | 0.079 | 0.084 |
| The whole of Japan | Males | 0–19 | Diabetes | 0.002 | 0.002 | 0.002 | 0.002 | 0.002 | 0.003 |
| The whole of Japan | Males | 20–39 | Diabetes | 0.018 | 0.018 | 0.019 | 0.020 | 0.021 | 0.024 |
| The whole of Japan | Males | 40–64 | Diabetes | 0.141 | 0.146 | 0.146 | 0.147 | 0.150 | 0.160 |
| The whole of Japan | Males | 65–74 | Diabetes | 0.318 | 0.323 | 0.330 | 0.328 | 0.331 | 0.337 |
| The whole of Japan | Females | 0–39 | Diabetes | 0.009 | 0.009 | 0.009 | 0.009 | 0.010 | 0.010 |
| The whole of Japan | Females | 40–74 | Diabetes | 0.108 | 0.110 | 0.108 | 0.108 | 0.110 | 0.113 |
| The whole of Japan | Females | 0–74 | Diabetes | 0.048 | 0.049 | 0.048 | 0.049 | 0.049 | 0.051 |
| The whole of Japan | Females | 0–19 | Diabetes | 0.002 | 0.002 | 0.002 | 0.002 | 0.003 | 0.003 |
| The whole of Japan | Females | 20–39 | Diabetes | 0.015 | 0.016 | 0.016 | 0.016 | 0.017 | 0.017 |
| The whole of Japan | Females | 40–64 | Diabetes | 0.076 | 0.079 | 0.078 | 0.077 | 0.079 | 0.084 |
| The whole of Japan | Females | 65–74 | Diabetes | 0.257 | 0.254 | 0.251 | 0.254 | 0.252 | 0.252 |
| The whole of Japan | Males and females | 0–39 | Mental disorders | 0.068 | 0.071 | 0.073 | 0.077 | 0.080 | 0.086 |
| The whole of Japan | Males and females | 40–74 | Mental disorders | 0.113 | 0.113 | 0.113 | 0.113 | 0.113 | 0.117 |
| The whole of Japan | Males and females | 0–74 | Mental disorders | 0.086 | 0.088 | 0.089 | 0.091 | 0.093 | 0.098 |
| The whole of Japan | Males | 0–39 | Mental disorders | 0.073 | 0.076 | 0.078 | 0.082 | 0.084 | 0.091 |
| The whole of Japan | Males | 40–74 | Mental disorders | 0.100 | 0.102 | 0.102 | 0.102 | 0.102 | 0.105 |
| The whole of Japan | Males | 0–74 | Mental disorders | 0.084 | 0.086 | 0.088 | 0.090 | 0.091 | 0.097 |
| The whole of Japan | Males | 0–19 | Mental disorders | 0.064 | 0.067 | 0.071 | 0.075 | 0.079 | 0.083 |
| The whole of Japan | Males | 20–39 | Mental disorders | 0.081 | 0.084 | 0.085 | 0.088 | 0.089 | 0.098 |
| The whole of Japan | Males | 40–64 | Mental disorders | 0.101 | 0.104 | 0.104 | 0.105 | 0.104 | 0.109 |
| The whole of Japan | Males | 65–74 | Mental disorders | 0.095 | 0.092 | 0.092 | 0.090 | 0.088 | 0.088 |
| The whole of Japan | Females | 0–39 | Mental disorders | 0.063 | 0.066 | 0.068 | 0.072 | 0.075 | 0.080 |
| The whole of Japan | Females | 40–74 | Mental disorders | 0.127 | 0.127 | 0.126 | 0.126 | 0.127 | 0.130 |
| The whole of Japan | Females | 0–74 | Mental disorders | 0.088 | 0.090 | 0.091 | 0.093 | 0.095 | 0.100 |
| The whole of Japan | Females | 0–19 | Mental disorders | 0.040 | 0.042 | 0.045 | 0.048 | 0.050 | 0.053 |
| The whole of Japan | Females | 20–39 | Mental disorders | 0.085 | 0.088 | 0.090 | 0.094 | 0.098 | 0.106 |
| The whole of Japan | Females | 40–64 | Mental disorders | 0.117 | 0.119 | 0.119 | 0.120 | 0.122 | 0.126 |
| The whole of Japan | Females | 65–74 | Mental disorders | 0.171 | 0.167 | 0.160 | 0.153 | 0.148 | 0.146 |
| Hokkaido Prefecture | Males | 0–39 | Hypertension | 0.009 | 0.011 | 0.011 | 0.012 | 0.012 | 0.012 |
| Aomori Prefecture | Males | 0–39 | Hypertension | 0.014 | 0.011 | 0.011 | 0.011 | 0.011 | 0.013 |
| Iwate Prefecture | Males | 0–39 | Hypertension | 0.012 | 0.011 | 0.010 | 0.012 | 0.013 | 0.013 |
| Miyagi Prefecture | Males | 0–39 | Hypertension | 0.012 | 0.012 | 0.011 | 0.012 | 0.012 | 0.012 |
| Akita Prefecture | Males | 0–39 | Hypertension | 0.015 | 0.011 | 0.010 | 0.012 | 0.013 | 0.012 |
| Yamagata Prefecture | Males | 0–39 | Hypertension | 0.011 | 0.011 | 0.015 | 0.016 | 0.017 | 0.015 |
| Fukushima Prefecture | Males | 0–39 | Hypertension | 0.010 | 0.010 | 0.011 | 0.012 | 0.013 | 0.012 |
| Ibaraki Prefecture | Males | 0–39 | Hypertension | 0.010 | 0.010 | 0.010 | 0.010 | 0.011 | 0.012 |
| Tochigi Prefecture | Males | 0–39 | Hypertension | 0.011 | 0.012 | 0.012 | 0.014 | 0.013 | 0.013 |
| Gunma Prefecture | Males | 0–39 | Hypertension | 0.012 | 0.010 | 0.009 | 0.010 | 0.010 | 0.011 |
| Saitama Prefecture | Males | 0–39 | Hypertension | 0.008 | 0.008 | 0.008 | 0.008 | 0.008 | 0.009 |
| Chiba Prefecture | Males | 0–39 | Hypertension | 0.009 | 0.009 | 0.009 | 0.010 | 0.009 | 0.009 |
| Tokyo Metropolitan Government | Males | 0–39 | Hypertension | 0.009 | 0.009 | 0.008 | 0.009 | 0.009 | 0.009 |
| Kanagawa Prefecture | Males | 0–39 | Hypertension | 0.009 | 0.008 | 0.009 | 0.009 | 0.009 | 0.009 |
| Niigata Prefecture | Males | 0–39 | Hypertension | 0.007 | 0.008 | 0.008 | 0.010 | 0.011 | 0.013 |
| Toyama Prefecture | Males | 0–39 | Hypertension | 0.012 | 0.010 | 0.012 | 0.012 | 0.011 | 0.013 |
| Ishikawa Prefecture | Males | 0–39 | Hypertension | 0.010 | 0.011 | 0.011 | 0.010 | 0.011 | 0.012 |
| Fukui Prefecture | Males | 0–39 | Hypertension | 0.009 | 0.008 | 0.008 | 0.010 | 0.011 | 0.013 |
| Yamanashi Prefecture | Males | 0–39 | Hypertension | 0.008 | 0.010 | 0.008 | 0.008 | 0.009 | 0.009 |
| Nagano Prefecture | Males | 0–39 | Hypertension | 0.011 | 0.012 | 0.011 | 0.012 | 0.012 | 0.012 |
| Gifu Prefecture | Males | 0–39 | Hypertension | 0.008 | 0.008 | 0.007 | 0.008 | 0.008 | 0.008 |
| Shizuoka Prefecture | Males | 0–39 | Hypertension | 0.010 | 0.010 | 0.010 | 0.010 | 0.010 | 0.010 |
| Aichi Prefecture | Males | 0–39 | Hypertension | 0.009 | 0.009 | 0.009 | 0.009 | 0.010 | 0.009 |
| Mie Prefecture | Males | 0–39 | Hypertension | 0.008 | 0.008 | 0.008 | 0.009 | 0.008 | 0.009 |
| Shiga Prefecture | Males | 0–39 | Hypertension | 0.011 | 0.008 | 0.009 | 0.009 | 0.009 | 0.010 |
| Kyoto Prefecture | Males | 0–39 | Hypertension | 0.008 | 0.008 | 0.007 | 0.007 | 0.008 | 0.008 |
| Osaka Prefecture | Males | 0–39 | Hypertension | 0.010 | 0.010 | 0.009 | 0.009 | 0.009 | 0.009 |
| Hyogo Prefecture | Males | 0–39 | Hypertension | 0.008 | 0.007 | 0.008 | 0.007 | 0.008 | 0.008 |
| Nara Prefecture | Males | 0–39 | Hypertension | 0.007 | 0.008 | 0.008 | 0.009 | 0.007 | 0.008 |
| Wakayama Prefecture | Males | 0–39 | Hypertension | 0.008 | 0.006 | 0.009 | 0.008 | 0.010 | 0.009 |
| Tottori Prefecture | Males | 0–39 | Hypertension | 0.010 | 0.008 | 0.007 | 0.009 | 0.009 | 0.011 |
| Shimane Prefecture | Males | 0–39 | Hypertension | 0.004 | 0.010 | 0.008 | 0.012 | 0.012 | 0.012 |
| Okayama Prefecture | Males | 0–39 | Hypertension | 0.009 | 0.009 | 0.009 | 0.009 | 0.010 | 0.010 |
| Hiroshima Prefecture | Males | 0–39 | Hypertension | 0.009 | 0.009 | 0.008 | 0.009 | 0.009 | 0.009 |
| Yamaguchi Prefecture | Males | 0–39 | Hypertension | 0.008 | 0.008 | 0.008 | 0.008 | 0.008 | 0.010 |
| Tokushima Prefecture | Males | 0–39 | Hypertension | 0.006 | 0.013 | 0.008 | 0.008 | 0.009 | 0.010 |
| Kagawa Prefecture | Males | 0–39 | Hypertension | 0.010 | 0.010 | 0.009 | 0.010 | 0.010 | 0.010 |
| Ehime Prefecture | Males | 0–39 | Hypertension | 0.007 | 0.008 | 0.007 | 0.009 | 0.008 | 0.009 |
| Kochi Prefecture | Males | 0–39 | Hypertension | 0.007 | 0.006 | 0.007 | 0.007 | 0.007 | 0.007 |
| Fukuoka Prefecture | Males | 0–39 | Hypertension | 0.008 | 0.009 | 0.009 | 0.009 | 0.010 | 0.009 |
| Saga Prefecture | Males | 0–39 | Hypertension | 0.010 | 0.010 | 0.010 | 0.008 | 0.008 | 0.010 |
| Nagasaki Prefecture | Males | 0–39 | Hypertension | 0.008 | 0.009 | 0.011 | 0.012 | 0.011 | 0.010 |
| Kumamoto Prefecture | Males | 0–39 | Hypertension | 0.008 | 0.009 | 0.010 | 0.010 | 0.011 | 0.011 |
| Oita Prefecture | Males | 0–39 | Hypertension | 0.010 | 0.011 | 0.009 | 0.009 | 0.010 | 0.009 |
| Miyazaki Prefecture | Males | 0–39 | Hypertension | 0.007 | 0.008 | 0.008 | 0.010 | 0.009 | 0.011 |
| Kagoshima Prefecture | Males | 0–39 | Hypertension | 0.010 | 0.008 | 0.008 | 0.008 | 0.009 | 0.010 |
| Okinawa Prefecture | Males | 0–39 | Hypertension | 0.009 | 0.015 | 0.012 | 0.012 | 0.013 | 0.013 |
| Hokkaido Prefecture | Males | 0–39 | Hyperlipidemia | 0.024 | 0.024 | 0.022 | 0.023 | 0.025 | 0.024 |
| Aomori Prefecture | Males | 0–39 | Hyperlipidemia | 0.023 | 0.021 | 0.021 | 0.020 | 0.024 | 0.023 |
| Iwate Prefecture | Males | 0–39 | Hyperlipidemia | 0.026 | 0.027 | 0.025 | 0.030 | 0.032 | 0.031 |
| Miyagi Prefecture | Males | 0–39 | Hyperlipidemia | 0.024 | 0.025 | 0.022 | 0.027 | 0.027 | 0.026 |
| Akita Prefecture | Males | 0–39 | Hyperlipidemia | 0.031 | 0.032 | 0.024 | 0.025 | 0.034 | 0.030 |
| Yamagata Prefecture | Males | 0–39 | Hyperlipidemia | 0.022 | 0.025 | 0.030 | 0.033 | 0.037 | 0.036 |
| Fukushima Prefecture | Males | 0–39 | Hyperlipidemia | 0.023 | 0.026 | 0.023 | 0.024 | 0.027 | 0.028 |
| Ibaraki Prefecture | Males | 0–39 | Hyperlipidemia | 0.023 | 0.022 | 0.021 | 0.020 | 0.021 | 0.021 |
| Tochigi Prefecture | Males | 0–39 | Hyperlipidemia | 0.032 | 0.030 | 0.027 | 0.028 | 0.028 | 0.029 |
| Gunma Prefecture | Males | 0–39 | Hyperlipidemia | 0.021 | 0.023 | 0.020 | 0.021 | 0.024 | 0.024 |
| Saitama Prefecture | Males | 0–39 | Hyperlipidemia | 0.016 | 0.014 | 0.013 | 0.015 | 0.016 | 0.016 |
| Chiba Prefecture | Males | 0–39 | Hyperlipidemia | 0.017 | 0.016 | 0.015 | 0.016 | 0.016 | 0.017 |
| Tokyo Metropolitan Government | Males | 0–39 | Hyperlipidemia | 0.017 | 0.018 | 0.016 | 0.017 | 0.018 | 0.019 |
| Kanagawa Prefecture | Males | 0–39 | Hyperlipidemia | 0.017 | 0.016 | 0.015 | 0.017 | 0.018 | 0.018 |
| Niigata Prefecture | Males | 0–39 | Hyperlipidemia | 0.022 | 0.024 | 0.023 | 0.026 | 0.030 | 0.031 |
| Toyama Prefecture | Males | 0–39 | Hyperlipidemia | 0.025 | 0.025 | 0.023 | 0.022 | 0.024 | 0.024 |
| Ishikawa Prefecture | Males | 0–39 | Hyperlipidemia | 0.021 | 0.019 | 0.017 | 0.018 | 0.022 | 0.022 |
| Fukui Prefecture | Males | 0–39 | Hyperlipidemia | 0.024 | 0.023 | 0.016 | 0.018 | 0.025 | 0.027 |
| Yamanashi Prefecture | Males | 0–39 | Hyperlipidemia | 0.017 | 0.019 | 0.016 | 0.017 | 0.016 | 0.015 |
| Nagano Prefecture | Males | 0–39 | Hyperlipidemia | 0.025 | 0.027 | 0.025 | 0.024 | 0.025 | 0.028 |
| Gifu Prefecture | Males | 0–39 | Hyperlipidemia | 0.015 | 0.016 | 0.014 | 0.017 | 0.019 | 0.020 |
| Shizuoka Prefecture | Males | 0–39 | Hyperlipidemia | 0.020 | 0.021 | 0.019 | 0.021 | 0.024 | 0.026 |
| Aichi Prefecture | Males | 0–39 | Hyperlipidemia | 0.018 | 0.019 | 0.019 | 0.020 | 0.022 | 0.023 |
| Mie Prefecture | Males | 0–39 | Hyperlipidemia | 0.014 | 0.014 | 0.014 | 0.018 | 0.018 | 0.018 |
| Shiga Prefecture | Males | 0–39 | Hyperlipidemia | 0.015 | 0.016 | 0.015 | 0.016 | 0.017 | 0.019 |
| Kyoto Prefecture | Males | 0–39 | Hyperlipidemia | 0.017 | 0.015 | 0.016 | 0.015 | 0.015 | 0.017 |
| Osaka Prefecture | Males | 0–39 | Hyperlipidemia | 0.018 | 0.018 | 0.017 | 0.017 | 0.017 | 0.017 |
| Hyogo Prefecture | Males | 0–39 | Hyperlipidemia | 0.016 | 0.014 | 0.014 | 0.015 | 0.018 | 0.018 |
| Nara Prefecture | Males | 0–39 | Hyperlipidemia | 0.014 | 0.014 | 0.011 | 0.013 | 0.013 | 0.014 |
| Wakayama Prefecture | Males | 0–39 | Hyperlipidemia | 0.016 | 0.017 | 0.018 | 0.016 | 0.016 | 0.017 |
| Tottori Prefecture | Males | 0–39 | Hyperlipidemia | 0.026 | 0.018 | 0.019 | 0.017 | 0.019 | 0.021 |
| Shimane Prefecture | Males | 0–39 | Hyperlipidemia | 0.014 | 0.020 | 0.022 | 0.024 | 0.027 | 0.026 |
| Okayama Prefecture | Males | 0–39 | Hyperlipidemia | 0.021 | 0.019 | 0.020 | 0.019 | 0.020 | 0.023 |
| Hiroshima Prefecture | Males | 0–39 | Hyperlipidemia | 0.017 | 0.018 | 0.017 | 0.018 | 0.019 | 0.019 |
| Yamaguchi Prefecture | Males | 0–39 | Hyperlipidemia | 0.013 | 0.013 | 0.012 | 0.015 | 0.017 | 0.018 |
| Tokushima Prefecture | Males | 0–39 | Hyperlipidemia | 0.016 | 0.022 | 0.014 | 0.020 | 0.018 | 0.019 |
| Kagawa Prefecture | Males | 0–39 | Hyperlipidemia | 0.017 | 0.019 | 0.018 | 0.018 | 0.022 | 0.022 |
| Ehime Prefecture | Males | 0–39 | Hyperlipidemia | 0.016 | 0.017 | 0.017 | 0.017 | 0.020 | 0.022 |
| Kochi Prefecture | Males | 0–39 | Hyperlipidemia | 0.018 | 0.011 | 0.012 | 0.015 | 0.016 | 0.017 |
| Fukuoka Prefecture | Males | 0–39 | Hyperlipidemia | 0.015 | 0.017 | 0.015 | 0.018 | 0.019 | 0.019 |
| Saga Prefecture | Males | 0–39 | Hyperlipidemia | 0.034 | 0.025 | 0.023 | 0.027 | 0.025 | 0.025 |
| Nagasaki Prefecture | Males | 0–39 | Hyperlipidemia | 0.014 | 0.015 | 0.016 | 0.019 | 0.019 | 0.021 |
| Kumamoto Prefecture | Males | 0–39 | Hyperlipidemia | 0.019 | 0.021 | 0.020 | 0.019 | 0.022 | 0.022 |
| Oita Prefecture | Males | 0–39 | Hyperlipidemia | 0.018 | 0.014 | 0.016 | 0.019 | 0.020 | 0.021 |
| Miyazaki Prefecture | Males | 0–39 | Hyperlipidemia | 0.016 | 0.015 | 0.017 | 0.022 | 0.022 | 0.025 |
| Kagoshima Prefecture | Males | 0–39 | Hyperlipidemia | 0.021 | 0.018 | 0.016 | 0.017 | 0.022 | 0.020 |
| Okinawa Prefecture | Males | 0–39 | Hyperlipidemia | 0.016 | 0.017 | 0.016 | 0.017 | 0.018 | 0.017 |
| Hokkaido Prefecture | Males | 0–39 | Diabetes | 0.013 | 0.013 | 0.013 | 0.014 | 0.014 | 0.013 |
| Aomori Prefecture | Males | 0–39 | Diabetes | 0.009 | 0.009 | 0.011 | 0.010 | 0.012 | 0.013 |
| Iwate Prefecture | Males | 0–39 | Diabetes | 0.007 | 0.006 | 0.008 | 0.008 | 0.008 | 0.009 |
| Miyagi Prefecture | Males | 0–39 | Diabetes | 0.010 | 0.011 | 0.010 | 0.011 | 0.010 | 0.010 |
| Akita Prefecture | Males | 0–39 | Diabetes | 0.009 | 0.009 | 0.008 | 0.008 | 0.010 | 0.010 |
| Yamagata Prefecture | Males | 0–39 | Diabetes | 0.010 | 0.010 | 0.010 | 0.013 | 0.015 | 0.013 |
| Fukushima Prefecture | Males | 0–39 | Diabetes | 0.006 | 0.007 | 0.007 | 0.008 | 0.009 | 0.009 |
| Ibaraki Prefecture | Males | 0–39 | Diabetes | 0.008 | 0.009 | 0.010 | 0.009 | 0.010 | 0.010 |
| Tochigi Prefecture | Males | 0–39 | Diabetes | 0.009 | 0.010 | 0.010 | 0.011 | 0.010 | 0.012 |
| Gunma Prefecture | Males | 0–39 | Diabetes | 0.008 | 0.009 | 0.009 | 0.011 | 0.010 | 0.010 |
| Saitama Prefecture | Males | 0–39 | Diabetes | 0.006 | 0.007 | 0.007 | 0.007 | 0.007 | 0.008 |
| Chiba Prefecture | Males | 0–39 | Diabetes | 0.009 | 0.009 | 0.008 | 0.009 | 0.009 | 0.009 |
| Tokyo Metropolitan Government | Males | 0–39 | Diabetes | 0.008 | 0.008 | 0.009 | 0.009 | 0.009 | 0.010 |
| Kanagawa Prefecture | Males | 0–39 | Diabetes | 0.007 | 0.007 | 0.007 | 0.009 | 0.009 | 0.009 |
| Niigata Prefecture | Males | 0–39 | Diabetes | 0.006 | 0.006 | 0.008 | 0.009 | 0.010 | 0.010 |
| Toyama Prefecture | Males | 0–39 | Diabetes | 0.010 | 0.010 | 0.010 | 0.009 | 0.010 | 0.010 |
| Ishikawa Prefecture | Males | 0–39 | Diabetes | 0.008 | 0.010 | 0.008 | 0.009 | 0.011 | 0.011 |
| Fukui Prefecture | Males | 0–39 | Diabetes | 0.006 | 0.008 | 0.007 | 0.009 | 0.010 | 0.011 |
| Yamanashi Prefecture | Males | 0–39 | Diabetes | 0.008 | 0.008 | 0.008 | 0.009 | 0.009 | 0.009 |
| Nagano Prefecture | Males | 0–39 | Diabetes | 0.008 | 0.007 | 0.008 | 0.008 | 0.008 | 0.008 |
| Gifu Prefecture | Males | 0–39 | Diabetes | 0.007 | 0.007 | 0.007 | 0.008 | 0.008 | 0.008 |
| Shizuoka Prefecture | Males | 0–39 | Diabetes | 0.009 | 0.009 | 0.009 | 0.010 | 0.011 | 0.011 |
| Aichi Prefecture | Males | 0–39 | Diabetes | 0.007 | 0.007 | 0.008 | 0.008 | 0.009 | 0.009 |
| Mie Prefecture | Males | 0–39 | Diabetes | 0.007 | 0.006 | 0.007 | 0.008 | 0.008 | 0.008 |
| Shiga Prefecture | Males | 0–39 | Diabetes | 0.007 | 0.008 | 0.007 | 0.007 | 0.009 | 0.009 |
| Kyoto Prefecture | Males | 0–39 | Diabetes | 0.006 | 0.006 | 0.006 | 0.006 | 0.007 | 0.007 |
| Osaka Prefecture | Males | 0–39 | Diabetes | 0.008 | 0.009 | 0.008 | 0.009 | 0.009 | 0.009 |
| Hyogo Prefecture | Males | 0–39 | Diabetes | 0.007 | 0.007 | 0.008 | 0.008 | 0.009 | 0.008 |
| Nara Prefecture | Males | 0–39 | Diabetes | 0.005 | 0.004 | 0.006 | 0.006 | 0.007 | 0.007 |
| Wakayama Prefecture | Males | 0–39 | Diabetes | 0.006 | 0.006 | 0.008 | 0.007 | 0.007 | 0.008 |
| Tottori Prefecture | Males | 0–39 | Diabetes | 0.005 | 0.005 | 0.008 | 0.009 | 0.009 | 0.008 |
| Shimane Prefecture | Males | 0–39 | Diabetes | 0.007 | 0.009 | 0.008 | 0.009 | 0.010 | 0.011 |
| Okayama Prefecture | Males | 0–39 | Diabetes | 0.007 | 0.006 | 0.008 | 0.008 | 0.008 | 0.009 |
| Hiroshima Prefecture | Males | 0–39 | Diabetes | 0.010 | 0.011 | 0.012 | 0.011 | 0.012 | 0.012 |
| Yamaguchi Prefecture | Males | 0–39 | Diabetes | 0.007 | 0.007 | 0.007 | 0.007 | 0.009 | 0.009 |
| Tokushima Prefecture | Males | 0–39 | Diabetes | 0.007 | 0.011 | 0.008 | 0.011 | 0.010 | 0.012 |
| Kagawa Prefecture | Males | 0–39 | Diabetes | 0.008 | 0.009 | 0.011 | 0.010 | 0.010 | 0.009 |
| Ehime Prefecture | Males | 0–39 | Diabetes | 0.005 | 0.007 | 0.007 | 0.008 | 0.009 | 0.010 |
| Kochi Prefecture | Males | 0–39 | Diabetes | 0.005 | 0.004 | 0.004 | 0.007 | 0.006 | 0.006 |
| Fukuoka Prefecture | Males | 0–39 | Diabetes | 0.007 | 0.007 | 0.008 | 0.008 | 0.009 | 0.009 |
| Saga Prefecture | Males | 0–39 | Diabetes | 0.006 | 0.005 | 0.005 | 0.007 | 0.006 | 0.007 |
| Nagasaki Prefecture | Males | 0–39 | Diabetes | 0.008 | 0.008 | 0.009 | 0.010 | 0.010 | 0.013 |
| Kumamoto Prefecture | Males | 0–39 | Diabetes | 0.008 | 0.008 | 0.008 | 0.009 | 0.009 | 0.010 |
| Oita Prefecture | Males | 0–39 | Diabetes | 0.008 | 0.008 | 0.007 | 0.009 | 0.011 | 0.010 |
| Miyazaki Prefecture | Males | 0–39 | Diabetes | 0.005 | 0.007 | 0.010 | 0.012 | 0.011 | 0.010 |
| Kagoshima Prefecture | Males | 0–39 | Diabetes | 0.008 | 0.007 | 0.008 | 0.009 | 0.009 | 0.010 |
| Okinawa Prefecture | Males | 0–39 | Diabetes | 0.006 | 0.009 | 0.006 | 0.006 | 0.008 | 0.008 |
| Hokkaido Prefecture | Males | 0–39 | Mental disorders | 0.065 | 0.069 | 0.067 | 0.071 | 0.073 | 0.073 |
| Aomori Prefecture | Males | 0–39 | Mental disorders | 0.059 | 0.057 | 0.058 | 0.058 | 0.062 | 0.062 |
| Iwate Prefecture | Males | 0–39 | Mental disorders | 0.046 | 0.052 | 0.053 | 0.059 | 0.060 | 0.063 |
| Miyagi Prefecture | Males | 0–39 | Mental disorders | 0.061 | 0.062 | 0.061 | 0.064 | 0.065 | 0.066 |
| Akita Prefecture | Males | 0–39 | Mental disorders | 0.060 | 0.061 | 0.060 | 0.065 | 0.067 | 0.071 |
| Yamagata Prefecture | Males | 0–39 | Mental disorders | 0.054 | 0.058 | 0.056 | 0.063 | 0.062 | 0.064 |
| Fukushima Prefecture | Males | 0–39 | Mental disorders | 0.057 | 0.061 | 0.063 | 0.067 | 0.065 | 0.067 |
| Ibaraki Prefecture | Males | 0–39 | Mental disorders | 0.049 | 0.050 | 0.052 | 0.053 | 0.057 | 0.062 |
| Tochigi Prefecture | Males | 0–39 | Mental disorders | 0.060 | 0.062 | 0.063 | 0.068 | 0.070 | 0.069 |
| Gunma Prefecture | Males | 0–39 | Mental disorders | 0.048 | 0.051 | 0.055 | 0.060 | 0.062 | 0.060 |
| Saitama Prefecture | Males | 0–39 | Mental disorders | 0.050 | 0.052 | 0.054 | 0.057 | 0.059 | 0.059 |
| Chiba Prefecture | Males | 0–39 | Mental disorders | 0.050 | 0.054 | 0.054 | 0.057 | 0.060 | 0.060 |
| Tokyo Metropolitan Government | Males | 0–39 | Mental disorders | 0.070 | 0.073 | 0.072 | 0.077 | 0.080 | 0.078 |
| Kanagawa Prefecture | Males | 0–39 | Mental disorders | 0.057 | 0.059 | 0.061 | 0.065 | 0.068 | 0.068 |
| Niigata Prefecture | Males | 0–39 | Mental disorders | 0.047 | 0.057 | 0.060 | 0.064 | 0.069 | 0.073 |
| Toyama Prefecture | Males | 0–39 | Mental disorders | 0.058 | 0.057 | 0.061 | 0.061 | 0.067 | 0.068 |
| Ishikawa Prefecture | Males | 0–39 | Mental disorders | 0.049 | 0.051 | 0.058 | 0.057 | 0.057 | 0.062 |
| Fukui Prefecture | Males | 0–39 | Mental disorders | 0.049 | 0.054 | 0.049 | 0.053 | 0.057 | 0.069 |
| Yamanashi Prefecture | Males | 0–39 | Mental disorders | 0.054 | 0.058 | 0.057 | 0.056 | 0.065 | 0.063 |
| Nagano Prefecture | Males | 0–39 | Mental disorders | 0.070 | 0.075 | 0.075 | 0.079 | 0.083 | 0.086 |
| Gifu Prefecture | Males | 0–39 | Mental disorders | 0.056 | 0.057 | 0.059 | 0.063 | 0.065 | 0.064 |
| Shizuoka Prefecture | Males | 0–39 | Mental disorders | 0.062 | 0.065 | 0.066 | 0.070 | 0.071 | 0.071 |
| Aichi Prefecture | Males | 0–39 | Mental disorders | 0.064 | 0.065 | 0.066 | 0.068 | 0.072 | 0.070 |
| Mie Prefecture | Males | 0–39 | Mental disorders | 0.053 | 0.052 | 0.053 | 0.055 | 0.057 | 0.057 |
| Shiga Prefecture | Males | 0–39 | Mental disorders | 0.058 | 0.058 | 0.052 | 0.050 | 0.054 | 0.051 |
| Kyoto Prefecture | Males | 0–39 | Mental disorders | 0.051 | 0.049 | 0.048 | 0.054 | 0.057 | 0.058 |
| Osaka Prefecture | Males | 0–39 | Mental disorders | 0.056 | 0.061 | 0.061 | 0.065 | 0.068 | 0.067 |
| Hyogo Prefecture | Males | 0–39 | Mental disorders | 0.056 | 0.058 | 0.059 | 0.063 | 0.066 | 0.065 |
| Nara Prefecture | Males | 0–39 | Mental disorders | 0.051 | 0.056 | 0.059 | 0.062 | 0.063 | 0.068 |
| Wakayama Prefecture | Males | 0–39 | Mental disorders | 0.053 | 0.061 | 0.060 | 0.062 | 0.068 | 0.069 |
| Tottori Prefecture | Males | 0–39 | Mental disorders | 0.074 | 0.071 | 0.064 | 0.067 | 0.073 | 0.076 |
| Shimane Prefecture | Males | 0–39 | Mental disorders | 0.051 | 0.060 | 0.057 | 0.059 | 0.069 | 0.068 |
| Okayama Prefecture | Males | 0–39 | Mental disorders | 0.076 | 0.083 | 0.083 | 0.082 | 0.085 | 0.081 |
| Hiroshima Prefecture | Males | 0–39 | Mental disorders | 0.067 | 0.070 | 0.068 | 0.070 | 0.075 | 0.073 |
| Yamaguchi Prefecture | Males | 0–39 | Mental disorders | 0.054 | 0.056 | 0.058 | 0.062 | 0.061 | 0.066 |
| Tokushima Prefecture | Males | 0–39 | Mental disorders | 0.059 | 0.065 | 0.071 | 0.068 | 0.075 | 0.076 |
| Kagawa Prefecture | Males | 0–39 | Mental disorders | 0.057 | 0.069 | 0.068 | 0.073 | 0.073 | 0.072 |
| Ehime Prefecture | Males | 0–39 | Mental disorders | 0.044 | 0.051 | 0.048 | 0.056 | 0.057 | 0.058 |
| Kochi Prefecture | Males | 0–39 | Mental disorders | 0.045 | 0.046 | 0.050 | 0.052 | 0.061 | 0.059 |
| Fukuoka Prefecture | Males | 0–39 | Mental disorders | 0.052 | 0.055 | 0.055 | 0.060 | 0.062 | 0.062 |
| Saga Prefecture | Males | 0–39 | Mental disorders | 0.051 | 0.058 | 0.058 | 0.065 | 0.068 | 0.073 |
| Nagasaki Prefecture | Males | 0–39 | Mental disorders | 0.060 | 0.068 | 0.067 | 0.071 | 0.075 | 0.074 |
| Kumamoto Prefecture | Males | 0–39 | Mental disorders | 0.053 | 0.057 | 0.060 | 0.062 | 0.066 | 0.064 |
| Oita Prefecture | Males | 0–39 | Mental disorders | 0.054 | 0.057 | 0.059 | 0.060 | 0.067 | 0.066 |
| Miyazaki Prefecture | Males | 0–39 | Mental disorders | 0.053 | 0.055 | 0.052 | 0.055 | 0.057 | 0.060 |
| Kagoshima Prefecture | Males | 0–39 | Mental disorders | 0.049 | 0.050 | 0.049 | 0.054 | 0.052 | 0.053 |
| Okinawa Prefecture | Males | 0–39 | Mental disorders | 0.046 | 0.053 | 0.057 | 0.058 | 0.060 | 0.061 |

**Table S4.**
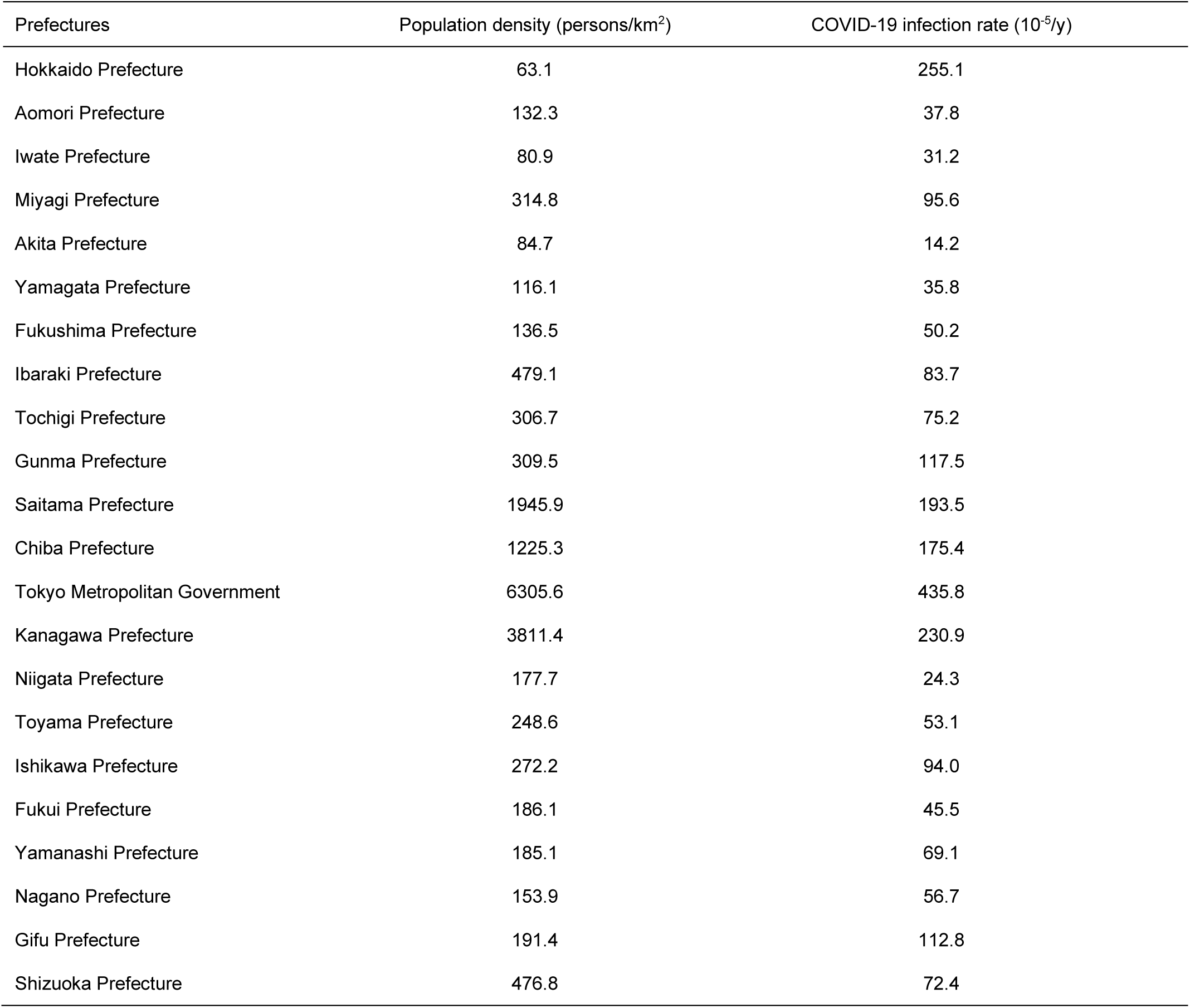
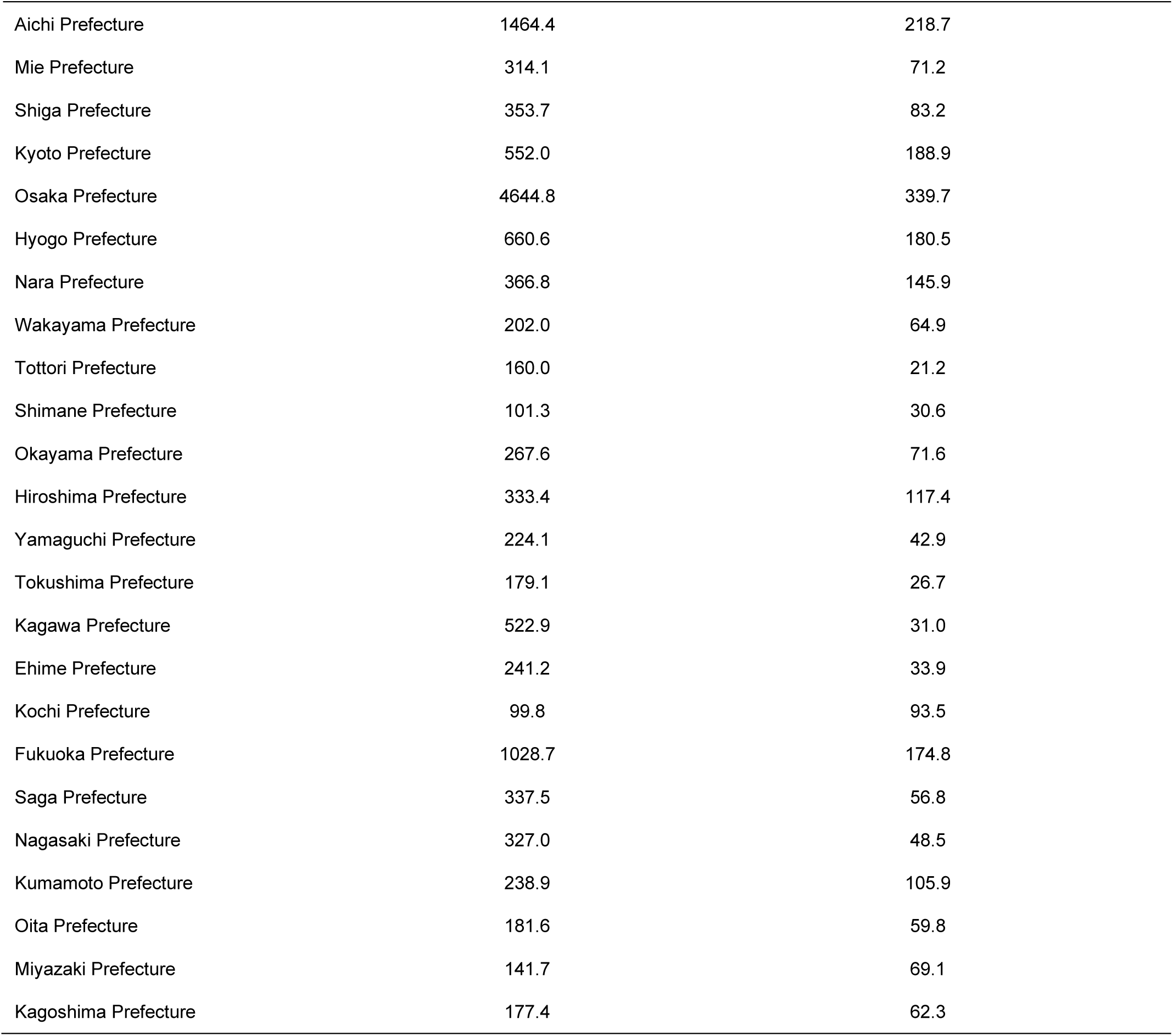
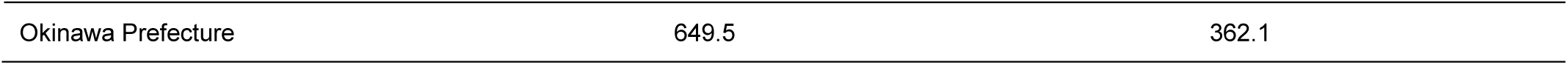
Population density and COVID–19 infection rate in 2020 in each prefecture.

| Prefectures | Population density (persons/km<sup>2</sup>) | COVID-19 infection rate (10<sup>-5</sup>/y) |
| --- | --- | --- |
| Hokkaido Prefecture | 63.1 | 255.1 |
| Aomori Prefecture | 132.3 | 37.8 |
| Iwate Prefecture | 80.9 | 31.2 |
| Miyagi Prefecture | 314.8 | 95.6 |
| Akita Prefecture | 84.7 | 14.2 |
| Yamagata Prefecture | 116.1 | 35.8 |
| Fukushima Prefecture | 136.5 | 50.2 |
| Ibaraki Prefecture | 479.1 | 83.7 |
| Tochigi Prefecture | 306.7 | 75.2 |
| Gunma Prefecture | 309.5 | 117.5 |
| Saitama Prefecture | 1945.9 | 193.5 |
| Chiba Prefecture | 1225.3 | 175.4 |
| Tokyo Metropolitan Government | 6305.6 | 435.8 |
| Kanagawa Prefecture | 3811.4 | 230.9 |
| Niigata Prefecture | 177.7 | 24.3 |
| Toyama Prefecture | 248.6 | 53.1 |
| Ishikawa Prefecture | 272.2 | 94.0 |
| Fukui Prefecture | 186.1 | 45.5 |
| Yamanashi Prefecture | 185.1 | 69.1 |
| Nagano Prefecture | 153.9 | 56.7 |
| Gifu Prefecture | 191.4 | 112.8 |
| Shizuoka Prefecture | 476.8 | 72.4 |
| Aichi Prefecture | 1464.4 | 218.7 |
| Mie Prefecture | 314.1 | 71.2 |
| Shiga Prefecture | 353.7 | 83.2 |
| Kyoto Prefecture | 552.0 | 188.9 |
| Osaka Prefecture | 4644.8 | 339.7 |
| Hyogo Prefecture | 660.6 | 180.5 |
| Nara Prefecture | 366.8 | 145.9 |
| Wakayama Prefecture | 202.0 | 64.9 |
| Tottori Prefecture | 160.0 | 21.2 |
| Shimane Prefecture | 101.3 | 30.6 |
| Okayama Prefecture | 267.6 | 71.6 |
| Hiroshima Prefecture | 333.4 | 117.4 |
| Yamaguchi Prefecture | 224.1 | 42.9 |
| Tokushima Prefecture | 179.1 | 26.7 |
| Kagawa Prefecture | 522.9 | 31.0 |
| Ehime Prefecture | 241.2 | 33.9 |
| Kochi Prefecture | 99.8 | 93.5 |
| Fukuoka Prefecture | 1028.7 | 174.8 |
| Saga Prefecture | 337.5 | 56.8 |
| Nagasaki Prefecture | 327.0 | 48.5 |
| Kumamoto Prefecture | 238.9 | 105.9 |
| Oita Prefecture | 181.6 | 59.8 |
| Miyazaki Prefecture | 141.7 | 69.1 |
| Kagoshima Prefecture | 177.4 | 62.3 |
| Okinawa Prefecture | 649.5 | 362.1 |

**Table S5.**
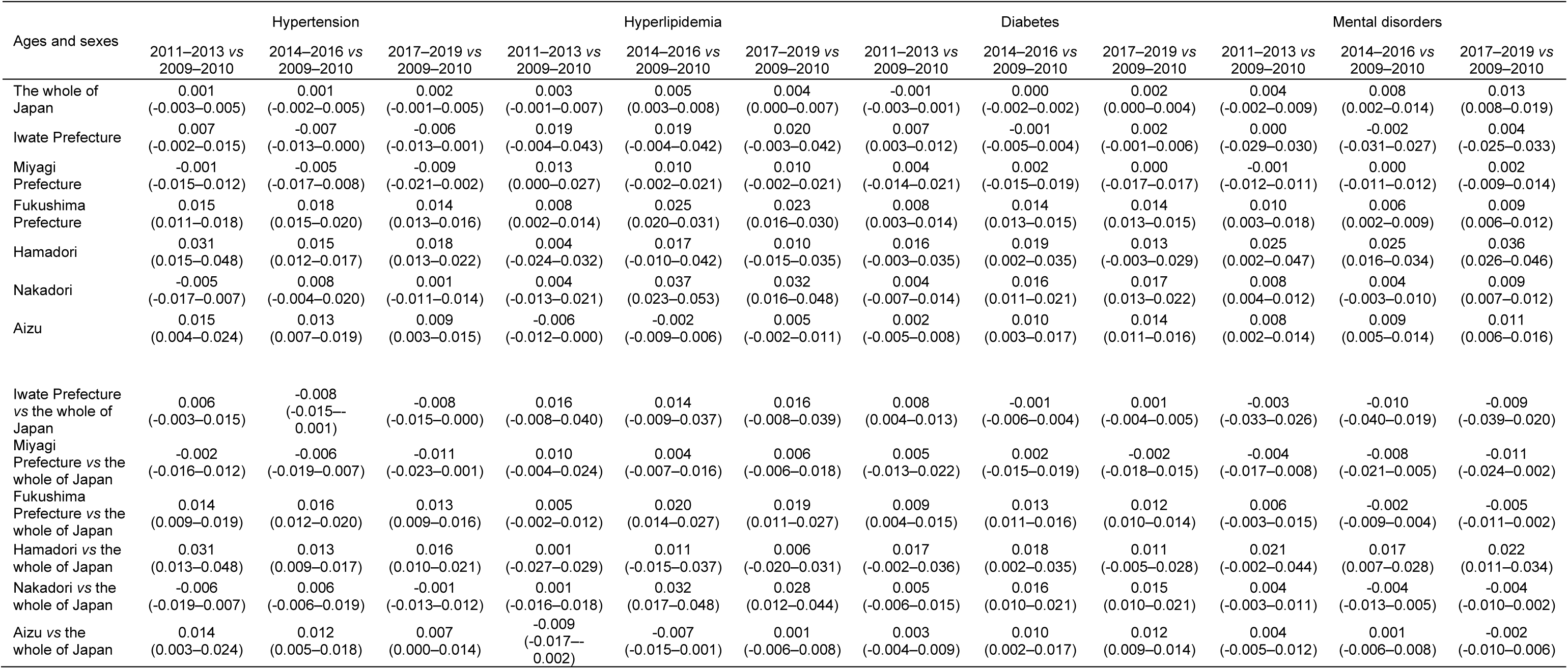
Difference in age-adjusted prevalence between three time periods and 2009–2010 (males and females, 0–74 years). Arithmetic mean (95% uncertainty interval).

**Table S6.**
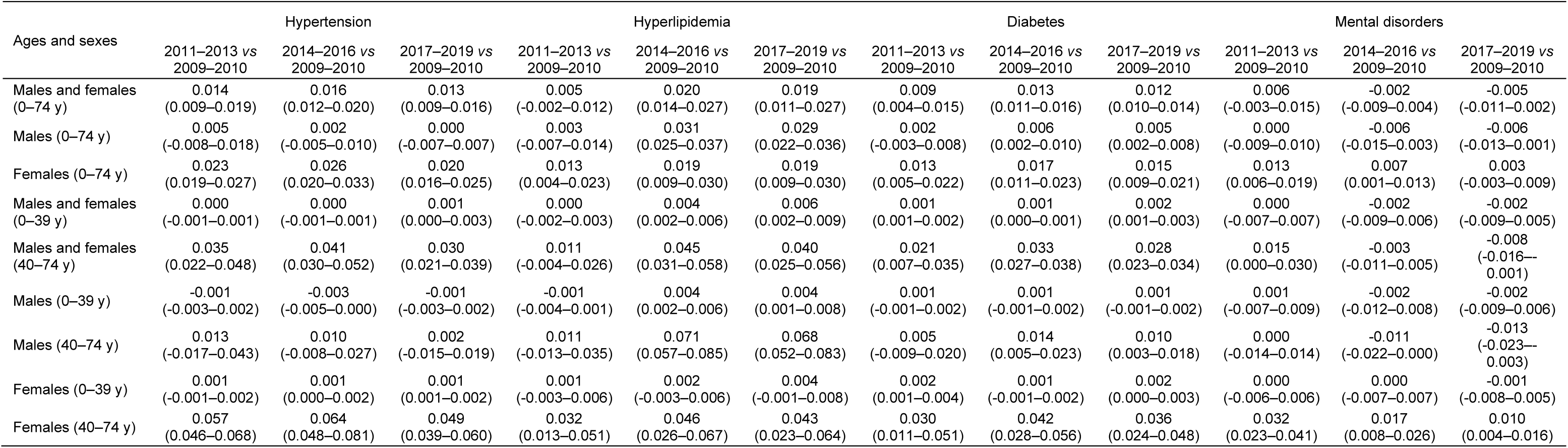
Difference in age-adjusted prevalence between three time periods and 2009–2010 by age groups and sex (Fukushima Prefecture vs the whole of Japan). Arithmetic mean (95% uncertainty interval).

**Table S7.**
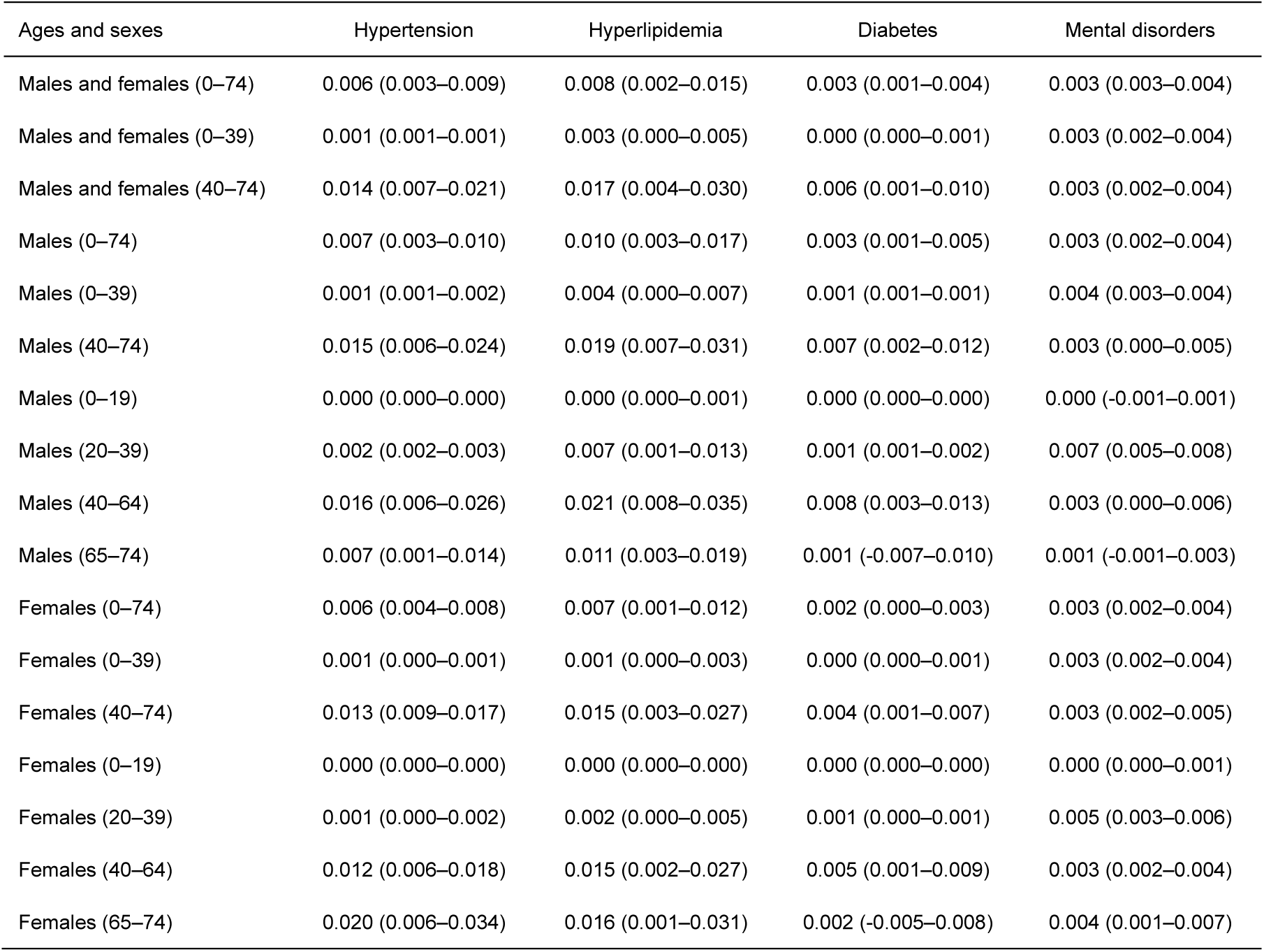
Difference between observed and estimated prevalence in 2020 by age groups and sex (the whole of Japan). Arithmetic mean (95% uncertainty interval).

**Table S8.**
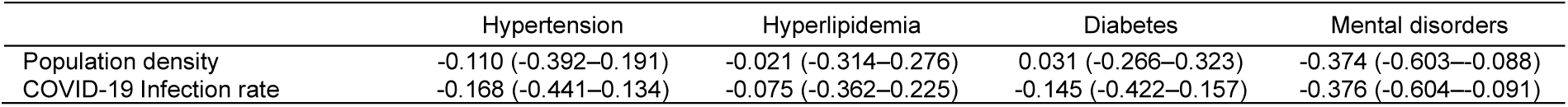
Correlations between population density or COVID-19 infection rate and ratio of observed to estimated age-adjusted prevalence of mental disorders in 2020 among 47 prefectures (males, 0–39 years). ρ: Spearman’s rank correlation coefficient (95% uncertainty interval).

**Table S9.**
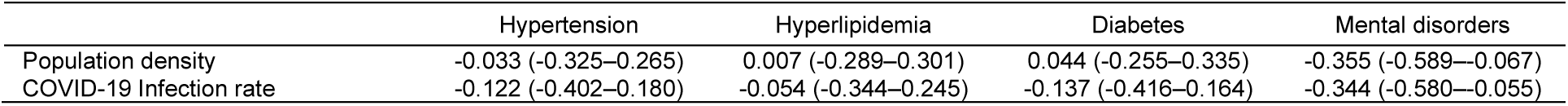
Correlations between population density or COVID-19 infection rate and difference between observed and estimated age-adjusted prevalence of mental disorders in 2020 among 47 prefectures (males, 0–39 years). ρ: Spearman’s rank correlation coefficient (95% uncertainty interval).

